# Optimal biannual COVID-19 vaccine boosting dates for those aged 65 and over

**DOI:** 10.1101/2025.11.20.25340687

**Authors:** Jeffrey P. Townsend, Hayley B. Hassler, Alex Dornburg

**Author notes:** Correspondence: Jeffrey P. Townsend, Department of Biostatistics, Yale School of Public Health; New Haven, Connecticut, USA, 06510-2483.

## Abstract

**Background:** To mitigate transmission and reduce the morbidity and mortality, the CDC recommends that individuals aged 65 and older receive a second dose of the 2024–2025 COVID-19 vaccine six months after their first booster dose. However, the optimal dates for biannual boosting are unknown. Geographic variation in infection rates throughout the year make it challenging to intuit the best yearly booster administration date to effectively prevent infection. With rapid waning and strong peaks associated with seasonality, precise timing of booster administration can maximize clinical and public health benefits.

**Methods:** We integrated longitudinal antibody waning trajectories, reinfection probabilities, and spatiotemporal projections of COVID-19 incidence to construct a geographically resolved model for optimizing booster timing in adults aged 65 and older.

**Results:** The benefit of dual booster vaccination is substantially increased by optimal timing, conferring as much as a 3–4-fold decrease in infection risk over five years. This benefit depends not only on the date of initial vaccination but also on the date of the second boost, which often falls outside of the current 6 month recommendation. In New York, for instance, optimal biannual booster dates are September 21 and February 22. In general, they are location-specific, relating to the geography of seasonal incidence.

**Conclusions:** Substantial benefit accrues from aptly timing dual COVID-19 booster vaccination campaigns for individuals over 65. Booster vaccination should be tailored to specific locations and anticipate regional periods of peak transmission. Analyses provide location-specific guidance for public health policy, healthcare provider recommendations, and individual decision-making.

**Summary:** Optimal timing for COVID-19 boosters improves their efficacy. Our analysis enables beneficial personalization of biannual booster schedules on the basis of infection history. This personalization improves public health.

**Key points:**

- Optimal biannual COVID-19 booster vaccination dates are in early autumn and late spring in most of the Northern Hemisphere.
- Optimal intervals between biannual boosts range from 4–7 months depending on location.
- Biannual boosting on the optimal dates provides up to 3–4 fold more protection.
- Delayed initial vaccination substantially alters the optimal second boost date.

## INTRODUCTION

Individuals aged 65 and older remain at disproportionate risk for hospitalization and death due to COVID-19, even as population-wide immunity has increased. This elevated vulnerability stems from age-associated declines in immune function, higher prevalence of comorbidities, and increased susceptibility to severe outcomes. To reduce morbidity and mortality in the aged, public health authorities have recommended biannual COVID-19 booster vaccinations [1] which have demonstrated substantial efficacy in preventing both infection [2] and severe disease [3], and have averted thousands of hospitalizations and deaths in older adults [4]. This protection wanes over time, necessitating timely reimmunization to sustain protection [5]. Indeed, there is a marked difference in antibody levels following vaccination in those aged 65 and older, resulting in substantially lower Anti-S IgG by 6 months [6]. The efficacy of a second booster at renewing protective antibody levels for these individuals is well-established. However, the benefit in terms of annual reduction in breakthrough infection as well as the optimal timing that would maximize that protection against breakthrough infections remains unclear. Alignment of booster administration with projected periods of elevated transmission has been shown to be highly beneficial for other seasonal viruses such as influenza [7], and could substantially improve vaccine-derived protection and reduce COVID-19 burden in this high-risk group.

Determination of the optimal timing for dual annual COVID-19 boosters is complicated by the evolving nature of SARS-CoV-2 seasonality. Initial efforts to define seasonal transmission patterns were hampered by pandemic-era dynamics, including shifting population behavior, waves driven by major antigenic shifts [8], and heterogeneous levels of immunological priming across age and exposure groups [9]. These features obscured the emergence of stable endemic seasonality. More recently, however, preliminary estimates of endemic-season transmission patterns based on phylogenetic ancestral and descendent states analysis [10] have provided a means to optimize the timing of single annual boosters [11] and could be applied to dual boosting as well.

Here we apply this approach. Integrating empirical antibody measurements from older adults within a comparative evolutionary framework, we estimated long-term waning rates and linked antibody levels to infection probability through a logistic model of consequent protection from infection. We then used comparative phylogenetic data on infection frequencies and seasonality to impute geographically specific daily probabilities of breakthrough infection across the year, incorporating the effects of all possible biannual pairs of boosts of antibody levels. This analysis enabled quantification of the long-term benefits of optimized biannual booster timing in older adults.

## METHODS

No long-term data is available on antibody waning following vaccination in older adults. Studies suggest similar short term (∼6 month) waning kinetics between older adult patients and younger adults [12,13]. However, the peak antibody responses are different between these groups ([12,13]). To model extended antibody trajectories in this population, we rescaled anti-Spike IgG values from Idda et al. ([14] Table 4) by adjusting their control group peak to align with empirically measured post-booster peak responses in a cohort of adults aged over 65 (*n* = 105; [12,13]). These values were then normalized to the peak antibody levels expected following BNT162b2 booster administration, enabling cross-cohort comparisons. Antibody decay profiles were reconstructed using a comparative phylogenetic framework applying ancestral and descendant state estimation to fit cohort-specific exponential waning parameters for SARS-CoV-2 [15,16], grounded in empirical data from endemic human coronaviruses including HCoV-OC43, NL63, and 229E [17]. This comparative evolutionary approach allows estimation of baseline post-infection antibody levels even in the absence of long-term SARS-CoV-2-specific data. Ancestral state reconstruction is a widely used method in evolutionary biology that leverages known trait distributions across a phylogeny to infer unknown states under a model of trait evolution [18–20]. We then used a linear model to relate anti-nucleocapsid and anti-spike titers, based on their strong empirical correlation, to further inform decay dynamics, as in Townsend et al. [21]. These baseline-adjusted profiles were then used to parameterize an augmented logistic waning model across treatment and control cohorts. Prior applications of this framework have demonstrated close agreement with empirical data [11,15], validating its utility in reconstructing antibody decay over extended timescales [22,23].

To convert modeled antibody trajectories into estimates of daily infection probability under endemic conditions, we leveraged the established link between circulating antibody levels and susceptibility to SARS-CoV-2 infection [24–27], a relationship that has been shown to be independent of symptom severity [28,29]. Indeed, the decline of anti-Spike IgG antibody levels is associated with increased susceptibility to reinfection [30], and they are an accurate correlate of protection against infection [31]. We used a logistic function to map daily antibody levels to daily probability of infection, parameterized by a slope and intercept representing the magnitude and inflection point of protection. These parameters were inferred jointly with baseline antibody levels using a phylogenetic comparative framework as in Townsend et al. [15].

To estimate relative monthly infection risk under endemic conditions, we used projections of seasonal COVID-19 incidence [10]. These projections were derived through a phylogenetic framework that reconstructed ancestral and descendant incidence states across human coronaviruses, drawing on long-term surveillance data from 12 Northern Hemisphere sites: Rochester (MN), New York City (NY), Edinburgh (UK), Stockholm (Sweden), Trøndelag (Norway), Gothenburg (Sweden), Amsterdam (Netherlands), South Korea (nationwide), Yamagata (Japan), Guangzhou (China), Sarlahi (Nepal), and Beersheba (Israel) [10]. Estimates proved consistent across alternative models of trait evolution and robust to different molecular phylogenies, time-calibrated chronograms, and non-recombinant sequence alignments. To assess the sensitivity of our results to these seasonality projections, we additionally evaluated the impact of deviations using observed 2024 incidence data.

Seasonal infection probabilities were based on long-term prevalence data from endemic human coronaviruses. These infection probabilities are the consequences of transmission patterns that have not been influenced by vaccination or public health interventions. However, in an endemic scenario with no interventions, an individual recently boosted against SARS-CoV-2 will experience lower risk of infection than an unboosted individual. To account for this, we adjusted the seasonal infection probabilities during the 30 days following booster administration by applying a time-resolved protection curve derived from clinical trial data of the BNT162b2 booster against the Delta variant (B.1.617.2) following primary series vaccination [32]. Daily protection levels were interpolated linearly from reported vaccine efficacy values at one and two weeks post-booster—peaking at one month. Beyond the first month, and continuing until the administration of a second booster dose, infection probabilities were attenuated using individualized estimates of antibody waning and corresponding reinfection risk [17].

Using these daily infection probabilities that account for antibody waning and the timing of both booster doses, we calculated cumulative breakthrough infection risk for all possible annual booster date pairings. For each boost, we assumed post-vaccination antibody responses as reported by Kometani et al. [13] and quantified subsequent benefit in terms of infection probabilities. Cumulative yearly risk was estimated by multiplying the probability of remaining uninfected through each preceding day by the probability of infection on the day of interest, repeated across the full annual cycle. These cumulative infection probabilities were computed for every day of the year and visualized for all booster date combinations using the plotly package in R. The paired booster dates associated with the lowest cumulative yearly risk were identified as the optimal biannual vaccination schedule.

We compared the relative benefit across seasons of optimal biannual boosting to the cases of no boosting or optimal annual boosting, using New York as our benchmark example. Our no boosting case was computed as the daily probability of infection of an individual at baseline antibody level year-round. Our optimal annual boosting case was calculated as the daily probability of infection of an elderly individual who was boosted on only the optimal day for annual boosting, and whose antibodies waned over the full year before the next boost [as in ref. 33 but specifying a typical antibody response to boosting for individuals aged >65]. Following Townsend et al. [17], booster administration was modeled to restore antibody levels to the empirically observed post-booster peak on the day of vaccination, as established in prior studies [34]. Following this peak, antibody decay was projected using a previously validated model of waning [16], which incorporates data-driven extrapolation beyond observed timepoints via phylogenetic ancestral and descendant states reconstruction [15]. Daily infection probabilities were computed by applying a logistic risk function, also derived from phylogenetic modeling, to the declining antibody levels, capturing the temporal erosion of protection due to both waning titers and antigenic drift in endemic SARS-CoV-2 variants [15]. These daily probabilities were then used to calculate cumulative risks of breakthrough infection across 6-month intervals over a period of five years. For each booster regimen, we calculated the cumulative probabilities of no breakthrough infections to estimate the long-term likelihood of remaining infection-free, accounting for the timing and frequency of scheduled boosts.

## RESULTS

As expected based on expected booster response, antibody waning, and endemic seasonality, biannual booster dates that are close together are inadvisable, and biannual booster dates with a greater interval between them are much better. Our analysis of New York shows a high range in benefit (14×) depending on the pair of dates selected for biannual boosting (**Fig. 1**), ranging from a probability of breakthrough infection of 2.5% for the worst pairing of dates to 0.017% for the best pairing. Optimal dates are September 21 and February 22 (**Fig. 1**). Indeed, biannual boosting on these optimal dates reduces the annual probability of infection in the elderly by 4.3-fold compared to a yearly boost on the optimal date [c.f. 11], substantially reducing the probability of breakthrough infection overall, but especially suppressing infections during the spring and summer as protection in the elderly wanes (**Fig. 2**). Over a five-year period, annual boosting provides substantial protection for the first six months after boosting, but breakthrough infection rates markedly increase subsequently until the next year’s booster. Optimally timed biannual boosting keeps the risk of infection uniformly low across the year, and yields a substantially lower cumulative probability of infection than annual boosting (**Fig. 2**).

**Figure 1.**
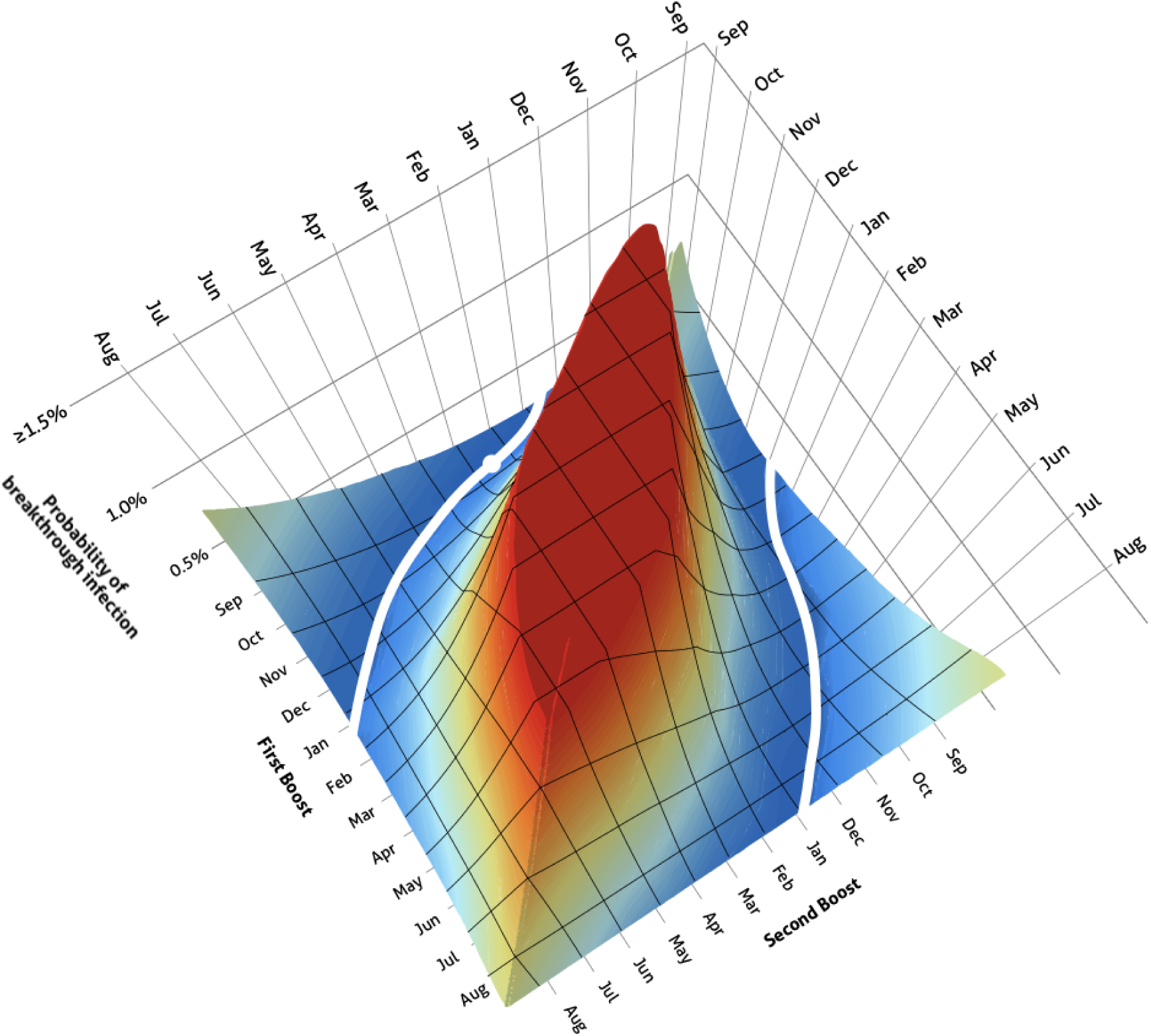
Cumulative probabilities of breakthrough infection over one year for each biannual boost pairing (high: red, low: blue; optimal second boost given first boost: white). A typical first boost in late September is optimally paired with a second boost in late February (white circle). ALT TEXT: Three-dimensional surface plot depicting the probability of breakthrough infection associated with each possible first and second booster date throughout the year, with lowest probabilities at substantial intervals between dates, and the lowest probability indicated for the date pair September 21 and February 22.

**Figure 2.**
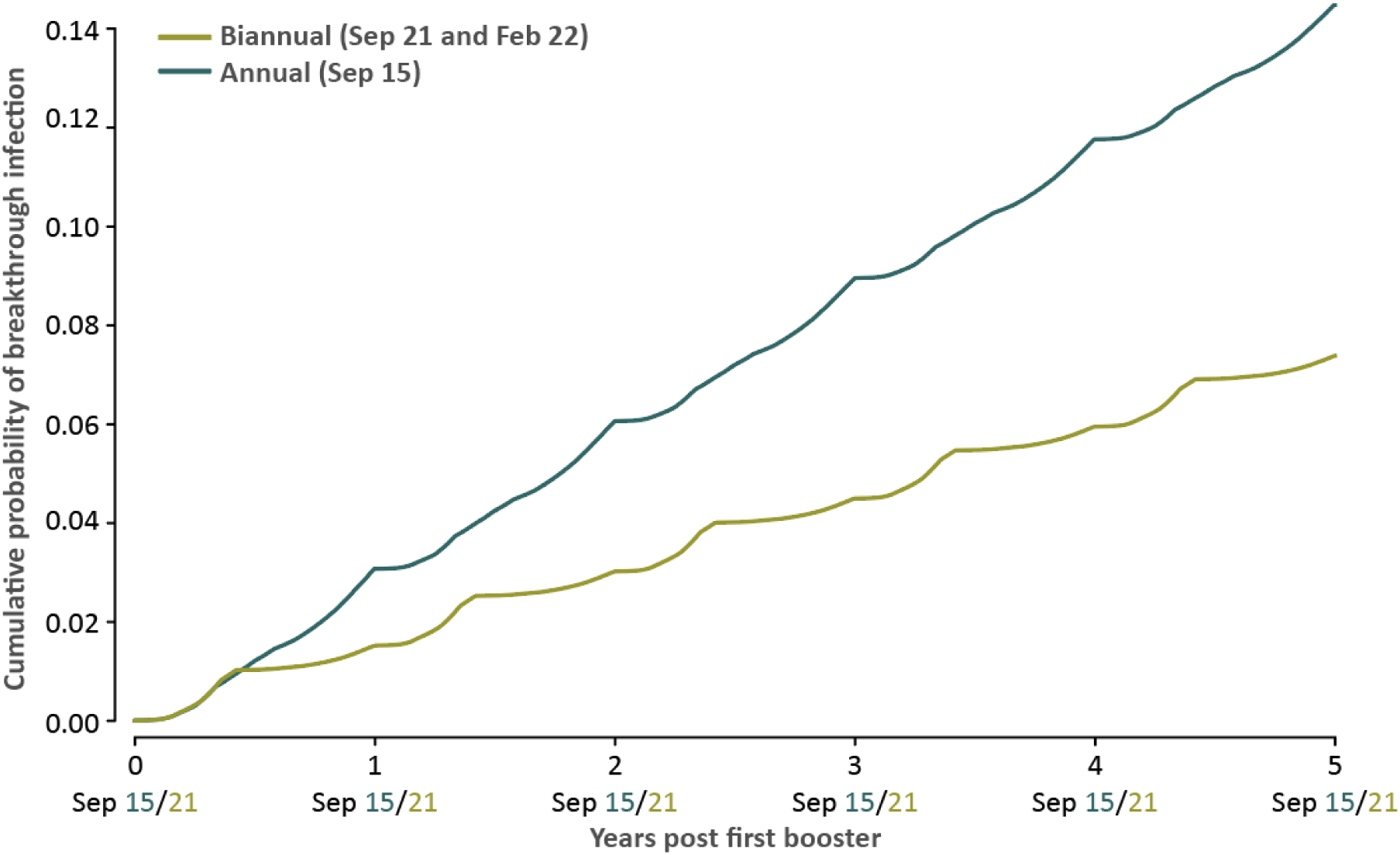
Cumulative probabilities of infection in New York over five years, with optimal annual boosting (dark teal) and with optimal biannual boosting (green). **ALT TEXT:** Plot of the cumulative probabilities of breakthrough infection over five years based on optimal annual and biannual boosting, providing evidence of substantially lower probabilities for optimal biannual boosting.

In other temperate locales of the Northern Hemisphere, the optimal dates for dual boosting are often similar to New York (**Supplementary Figs. 1 A–E; Supplementary Table 1 A–E**).

However, optimal dual booster dates for endemic COVID-19 can be as late as October / February and November / May (Sweden), December / July (Nepal, Japan), and even February / July (Israel; **Table 1**; Supplementary Figs. 1 A–E; Supplementary Tables 1 A–E). Across all locations, optimally timed biannual boosting keeps the risk of infection uniformly low across the year, and yields a substantially lower cumulative probability of infection than annual boosting (**Supplementary Fig. 2**), with a dramatic reduction in breakthrough infection risk for biannual versus annual boosting (**Supplementary Fig. 3**).

**Table 1.** Optimal biannual booster dates and mean yearly probabilities of breakthrough infection.

| Location | First booster | Second booster | Cumulative probability of breakthrough infection over one year |
| --- | --- | --- | --- |
| New York | September 21 | February 22 | 0.17% |
| Stockholm, Sweden | October 19 | February 21 | 0.13% |
| Guangzhou, China | September 24 | February 5 | 0.16% |
| Edinburgh, UK | September 20 | February 1 | 0.11% |
| Beersheba, Israel | February 28 | July 29 | 0.15% |
| Nepal | December 28 | July 31 | 0.19% |
| Amsterdam, Netherlands | September 28 | March 13 | 0.22% |
| Trondelag, Norway | September 24 | February 5 | 0.16% |
| South Korea | September 28 | February 19 | 0.15% |
| Gothenburg, Sweden | November 23 | May 15 | 0.17% |
| Yamagata, Japan | December 23 | July 3 | 0.16% |

The intervals between the optimal first and second dose were often divergent from the CDC-recommended 6-month delay between boosters. Intervals between boosters with an optimally timed first booster ranged from around four months (Stockholm, Sweden) to over seven months (Nepal; **Table 1**). These divergences from a 6-mocan have substantial consequences on breakthrough infection risks. Across all locations, optimal biannual booster dates yield substantially lower cumulative probabilities of breakthrough infections than other pairs of dates determined by six-month delays (**Fig. 3**). The extent of benefit of vaccinating on optimal dates in preference to vaccinating with a six-month delay varies substantially by location and by first booster date (**Fig. 3**). If a first booster is non-optimally timed, optimal dates for second boosters in all locations are even more divergent from the 6-month delay, especially for later first-boost dates (**Supplementary Table 1**).

**Figure 3.**
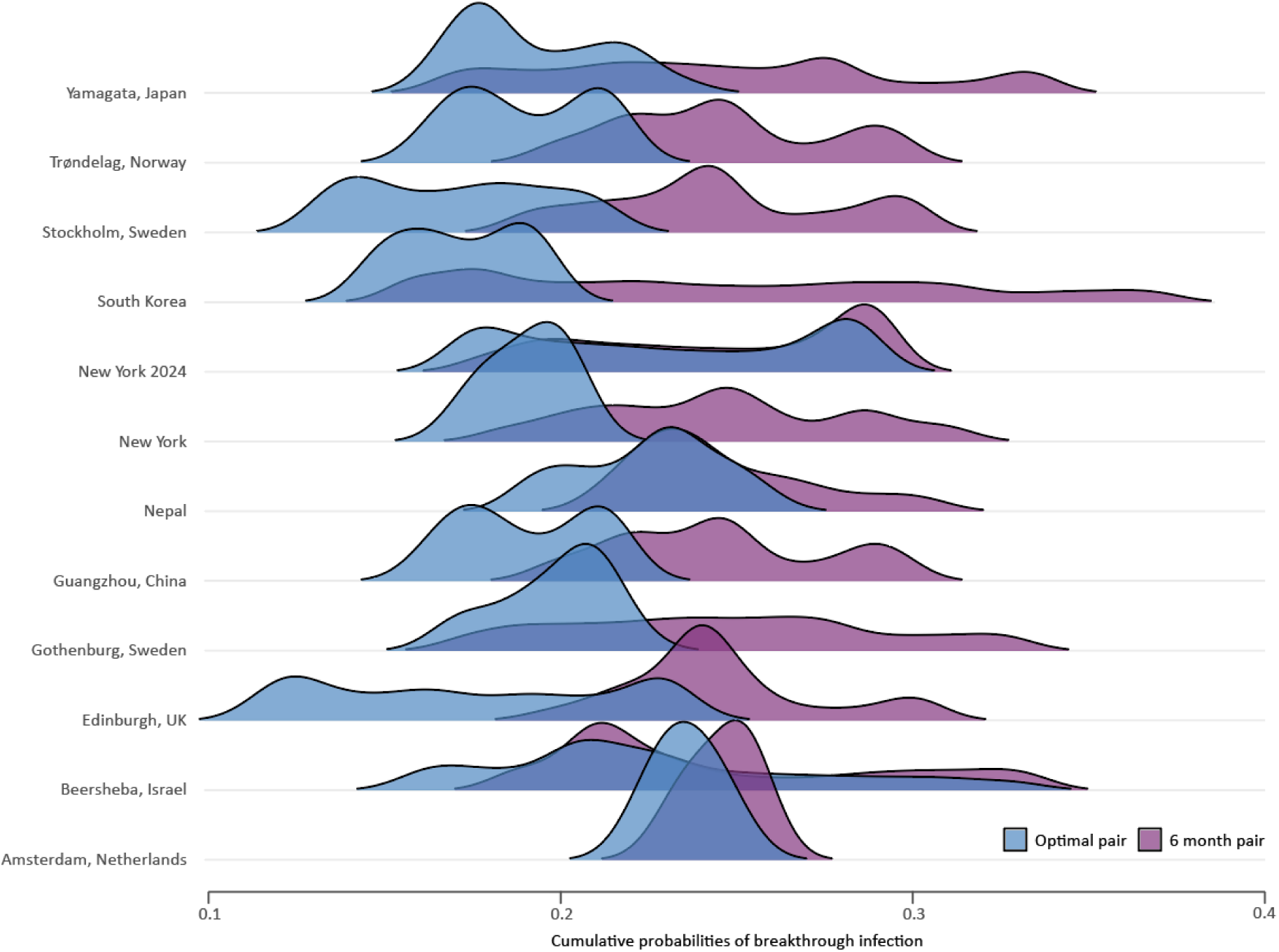
Distributions of cumulative probabilities of breakthrough infection over one year for optimal biannual booster date pairs (blue) compared to six-month booster pairs (purple) for twelve Northern Hemispheric locations. Given a first booster date, a second booster date always performs as well or better if determined by an optimal delay than by a six-month delay. **ALT TEXT:** Ridgeline plot illustrating that cumulative probabilities of breakthrough infection are substantially lower when boosters are administered at optimal biannual booster dates than when administered at fixed six-month intervals.

Our reanalysis of optimal dates for New York based on 2024 incidence instead of projected endemic incidence yielded modest differences in optimal date pairs (**Supplementary Fig. 1E**). Based on 2024 monthly incidence in New York, the optimal biannual date pair was September 30 for the first booster (one week later than based on endemic projections) and April 14 for the second booster (three weeks later than that based on endemic projections).

## DISCUSSION

Here, we have estimated that elderly individuals following an optimal dual annual booster strategy experience, on average, a 14-fold reduction in risk relative to those receiving a single optimally timed annual booster. Across geographic settings, the first booster is consistently optimized when administered in the fall through early winter, aligning with prior evidence suggesting enhanced effectiveness during this period [33]. However, the second booster does not universally align with the fixed six-month interval currently recommended in clinical guidance. Rather, the benefits gained by the timing of the second dose are influenced by the specific date of the first: for example, a September booster in New York corresponds well with a second dose in February (5 months), while an October first booster favors a delay into May (7 months). This modulation of optimal spacing between booster dates is also observed across other locations, driven by regional variation in seasonal incidence patterns.

The geographically specific and population-informed datasets we have used to estimate optimal booster vaccination timing may not fully capture short-term fluctuations and patterns in current transmission dynamics. For example, many regions have continued to experience summer and winter infection surges, reflecting the early transmission history of the COVID-19 pandemic. To address this issue, we explicitly modeled the impact of summer surges consistent with New York 2024 incidences on our outcomes. Such deviations modulate optimal timing to a limited extent. However, the broader structure of optimality remains intact. Moreover, we demonstrated that, in general, optimally determined booster pairs substantially outperform the CDC recommendation of a six-month delay [1]. In general, respiratory virus seasonality, even at endemicity, is not static. Peak periods of infection shift forward or backward and vary in magnitude across years [35–37]. In seasons where surges occur earlier than anticipated, initiating booster vaccination ahead of the predicted optimal window may confer substantially greater protection than adhering strictly to modeled dates. These findings underscore the value of setting data-informed quantitative expectations, but also adapting recommendations to evolving transmission landscapes.

Our analysis is based on data from individuals aged 65 and older with no ongoing severe medical conditions and no medications that affect immune function. There is clinically meaningful heterogeneity in comorbidities and in immune function within the 65 and older demographic. Older adults have the highest rates of prescription medication use across age groups [38], and several commonly prescribed drugs, including immunosuppressants, can diminish vaccine-induced immune responses. Prior studies have highlighted subgroups, such as patients undergoing B-cell–depleting therapies (e.g., rituximab) [39] or disease-modifying treatments for multiple sclerosis [40], for whom booster vaccine efficacy may be reduced, and for whom alternative boosting intervals may be warranted. These heterogeneities may affect optimal biannual booster timing [41]. However, the effect of these heterogeneities on optimal biannual boosting dates is likely small, because these heterogeneities generally affect the magnitude of protection provided more than the optimal timing of protection. The best timing for individuals with specific health concerns may be further resolved through clinician–patient discussions that evaluate individual risk profiles and the potential consequences of infection. Additionally, our model focuses on the probability of infection as predicted by antibody titer, an established correlate of protection [42]. We do not predict disease severity, as this requires integrating additional immune parameters, including T-cell function, inflammatory markers, and host-specific risk factors.

Our analysis assumes that both booster doses confer equivalent immune responses, but this assumption does not fully capture the nuances of booster effectiveness in practice. When booster doses are administered in close succession, particularly in unrealistic scenarios such as back-to-back vaccinations on February 1st and 2nd, the immunological benefit of the second dose would likely be substantially reduced due to antibody-mediated antigen clearance, wherein preexisting antibodies bind and neutralize a newly introduced antigen and it is removed before exposure to an antigen-presenting cells, limiting additional activation of naïve B cells and T cells and diminishing the efficacy of subsequent exposures or booster immunizations. Fortunately, such extremely short intervals between boosts are deemed highly inadvisable by our approach anyway. Moreover, such timing is not representative or realistic in clinical practice. Moderate levels of antibody-mediated antigen clearance may also diminish the net protective effect of some non-optimal timings, but are unlikely to affect optimal biannual booster dates and are likely to have minimal impact on the estimated protective effect.

A more salient consideration is that, as long as production of the biannual booster supply is performed on the basis of antigens identified at one time prior to the first boost, second booster doses will likely offer somewhat diminished protection compared to first booster doses, due to ongoing antigenic evolution of circulating strains that leads to increasing mismatch with a static vaccine design. Even so, second doses are unlikely to be grossly mismatched. In general, it may be prudent to schedule the second booster just slightly earlier to ensure sufficient protection through peak incidence, though doing so would likely only trivially affect the predicted outcomes. Further research on the degree of diminished efficacy of vaccines as molecular evolution of antigenic sites occurs would be warranted.

Our work represents the first continuous, quantitative assessment of risk reduction across all potential annual booster timing combinations in older adults. Our approach provides actionable timing recommendations that adapt to the first booster date, and frames these recommendations within a quantitative approach responsive to projected annual transmission patterns. This work provides a critical foundation for public health policy and clinical guidance by establishing timing strategies that maximize protection in the general elderly population that can be readily adapted as more data become available on specific therapies or immunological profiles. This framework can be applied to inform not only COVID-19 booster policy, but also policies for other seasonal respiratory viruses. Such insights enable improvements in booster effectiveness in older adults and support informed decision-making by clinicians and public health authorities.

## Supplementary Data

Supplementary Figures and Tables are available for this paper. Consisting of data provided by the authors to benefit the reader, the posted materials are not copyedited and are the sole responsibility of the authors, so questions or comments should be addressed to the corresponding author.

## Notes

### Author contributions

JPT, HBH, and AD conceived the project and designed the study; HBH performed formal analyses with guidance from JPT and AD; JPT, HBH, and AD designed data visualizations; HBH implemented data visualizations; AD and JPT wrote the manuscript; and all authors reviewed the manuscript before submission. JPT and AD were responsible for the decision to submit the manuscript. All authors had full access to all the data in the study and had final responsibility for the decision to submit for publication.

### Data availability

All data, imputed monthly proportions, and code underlying this study are publicly available on Zenodo: DOI:10.5281/zenodo.17238313.

### Potential conflicts of interest

The authors declare no conflicts of interest.

### Financial support

National Science Foundation of the United States of America RAPID 2031204 (JPT and AD), NSF Expeditions CCF 1918784 (JPT), and support from the University of North Carolina, Charlotte to AD.

## Notes

### Competing Interest Statement

The authors have declared no competing interest.

