## Supplementary Fig. 1 for "Optimal biannual COVID-19 vaccine boosting dates for those aged 65 and over"

**A**

### STOCKHOLM, SWEDEN

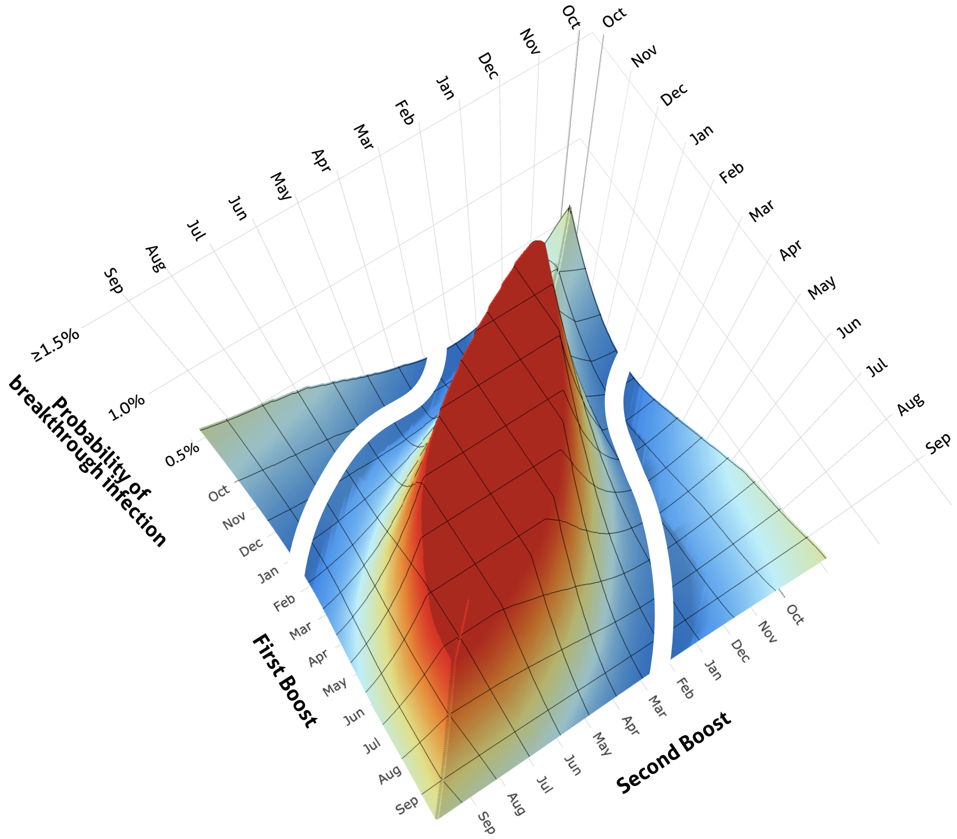


**B**

### GOTHENBURG, SWEDEN

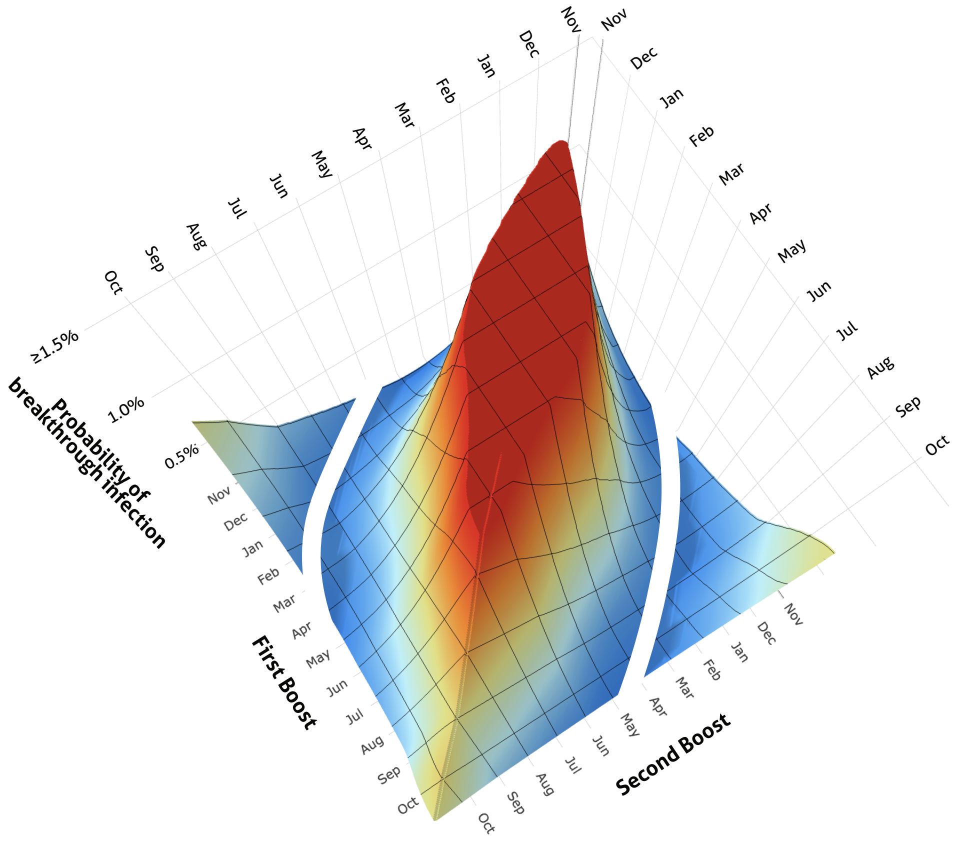

C

### NEPAL

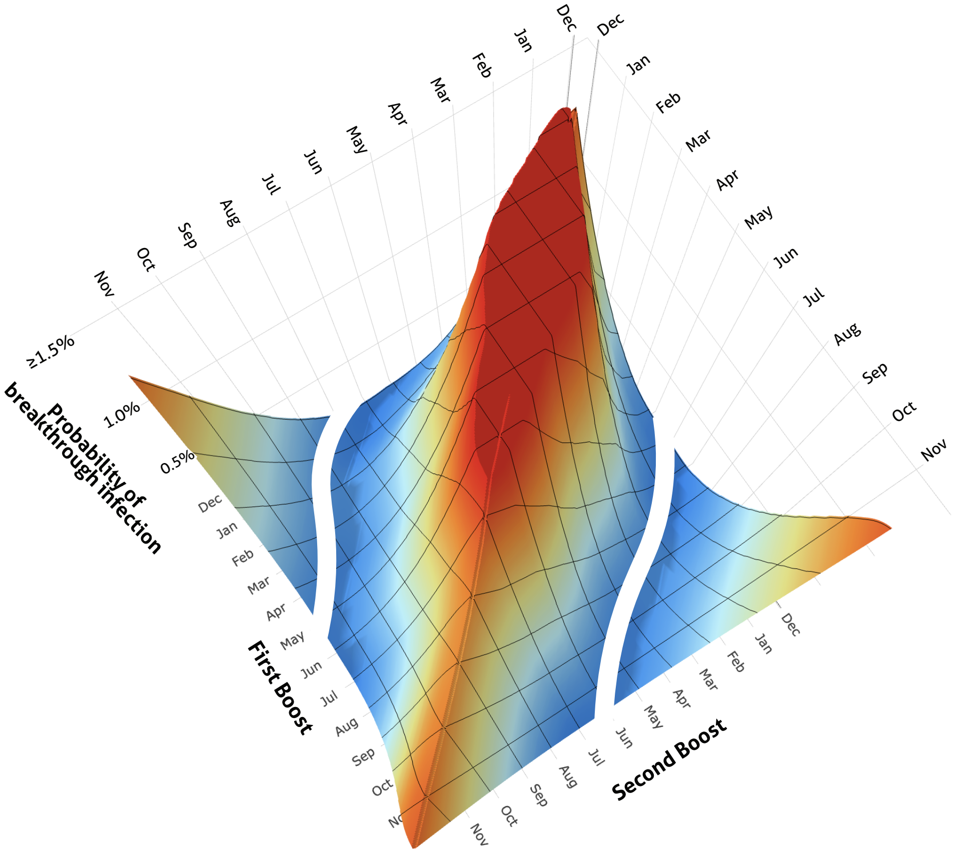


D

### BEERSHEBA, ISRAEL

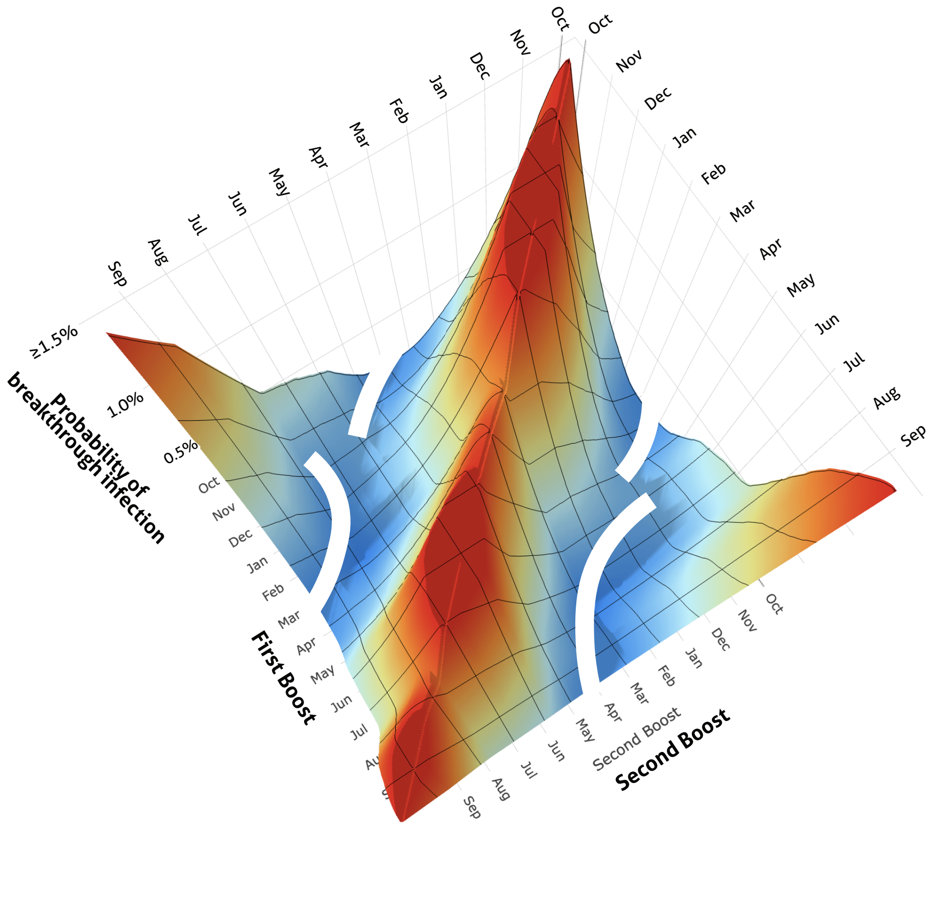

**E** NEW YORK CITY (2024)

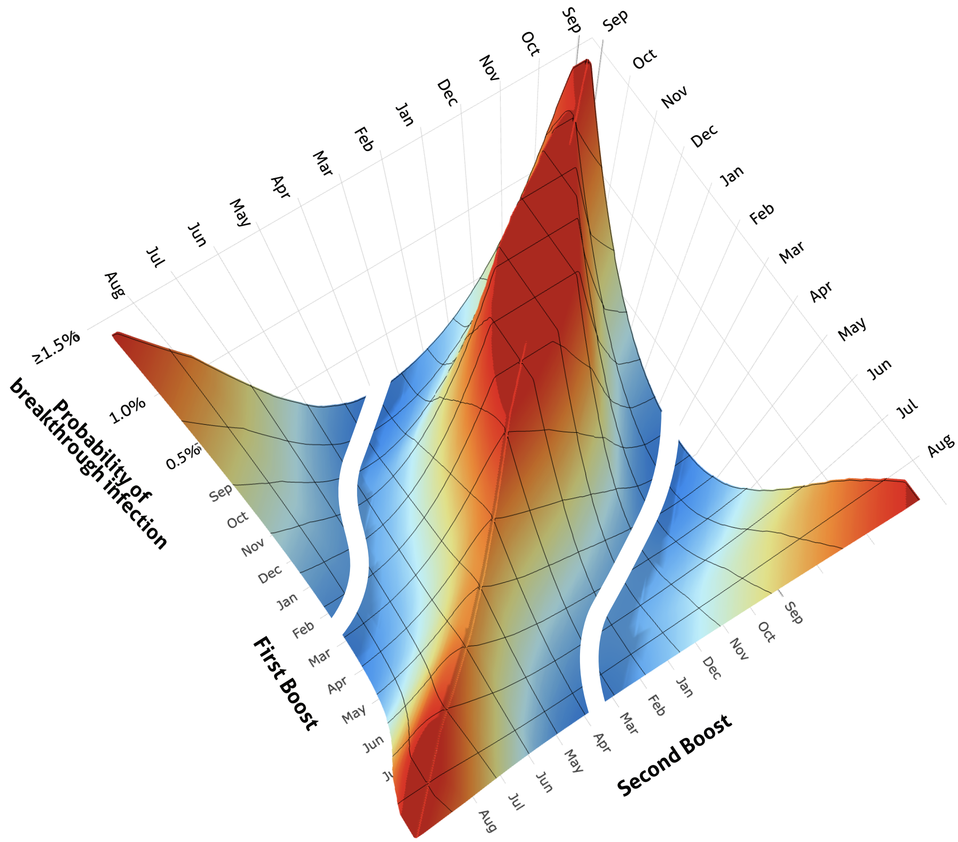


**F** TRONDELAG, NORWAY

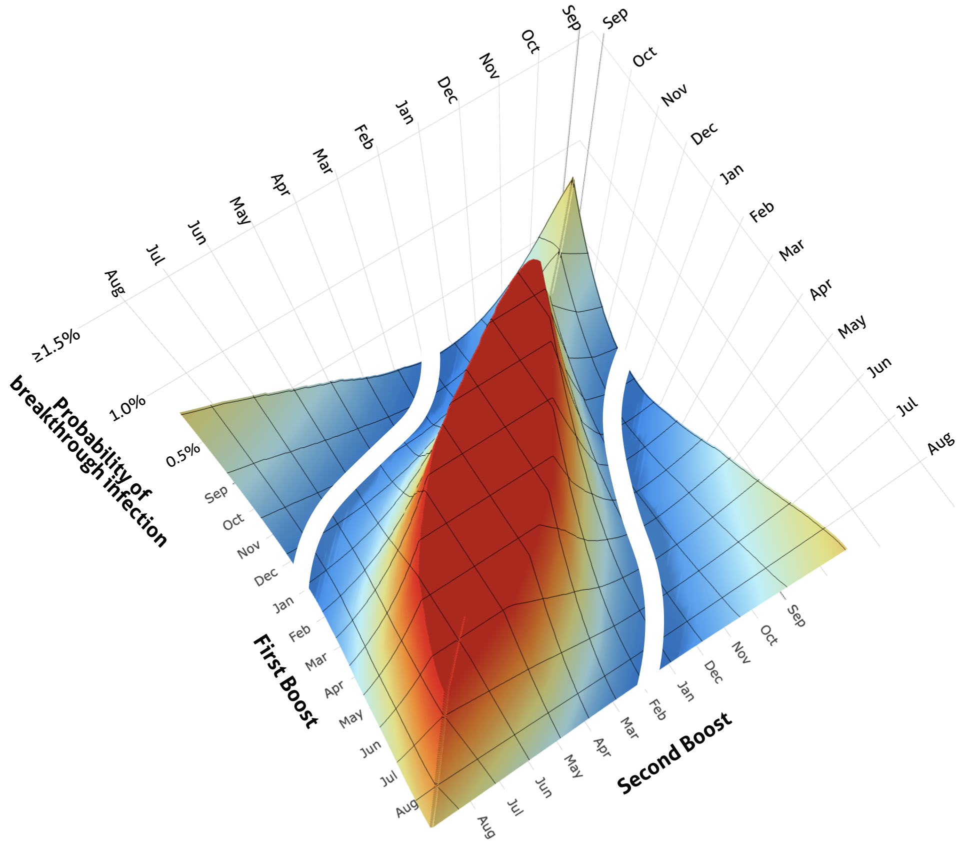

G

### AMSTERDAM, NETHERLANDS

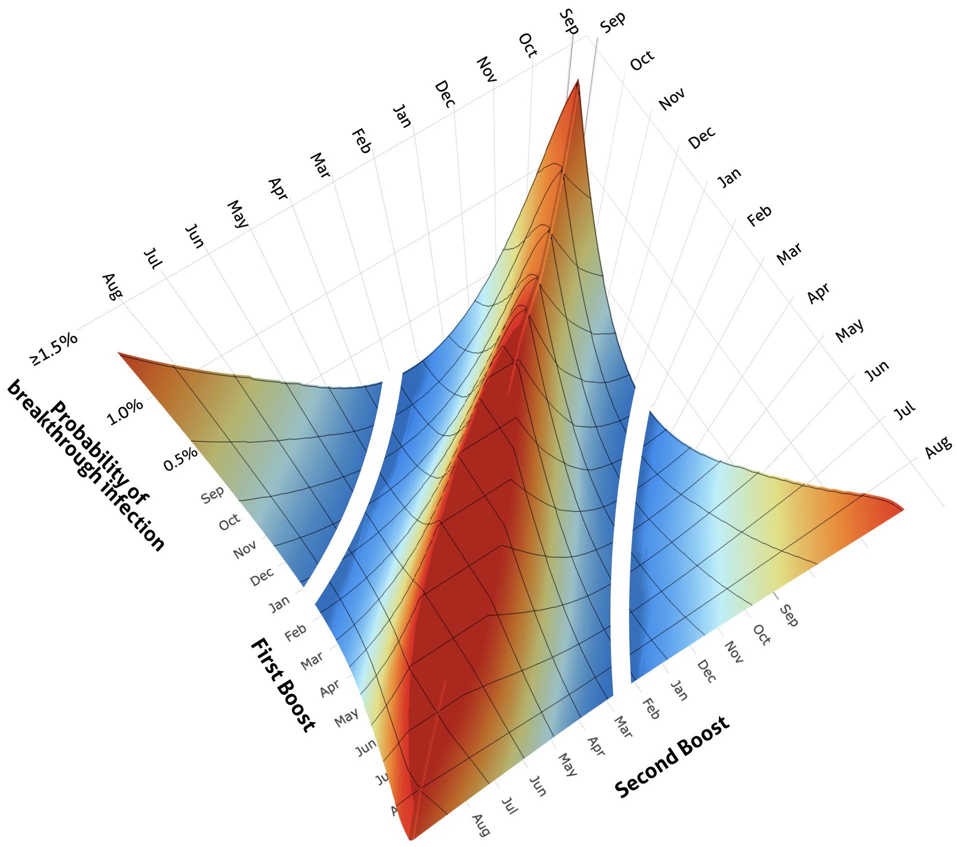


H

### SOUTH KOREA

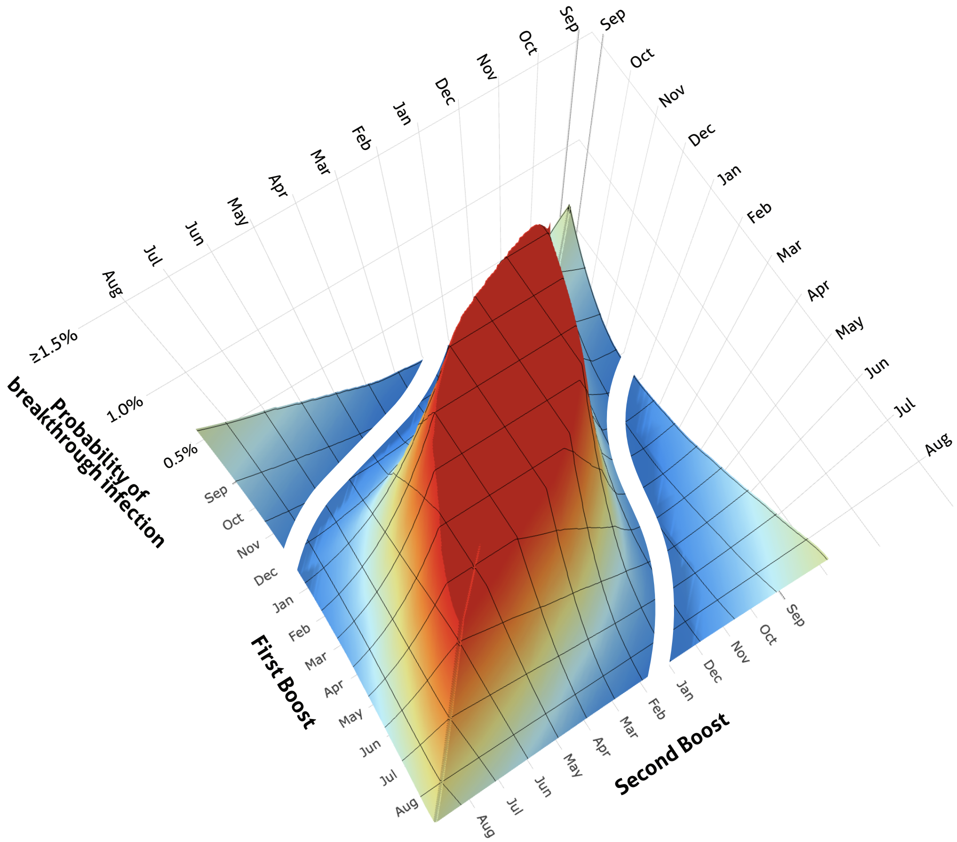

**I**

### EDINBURGH, UK

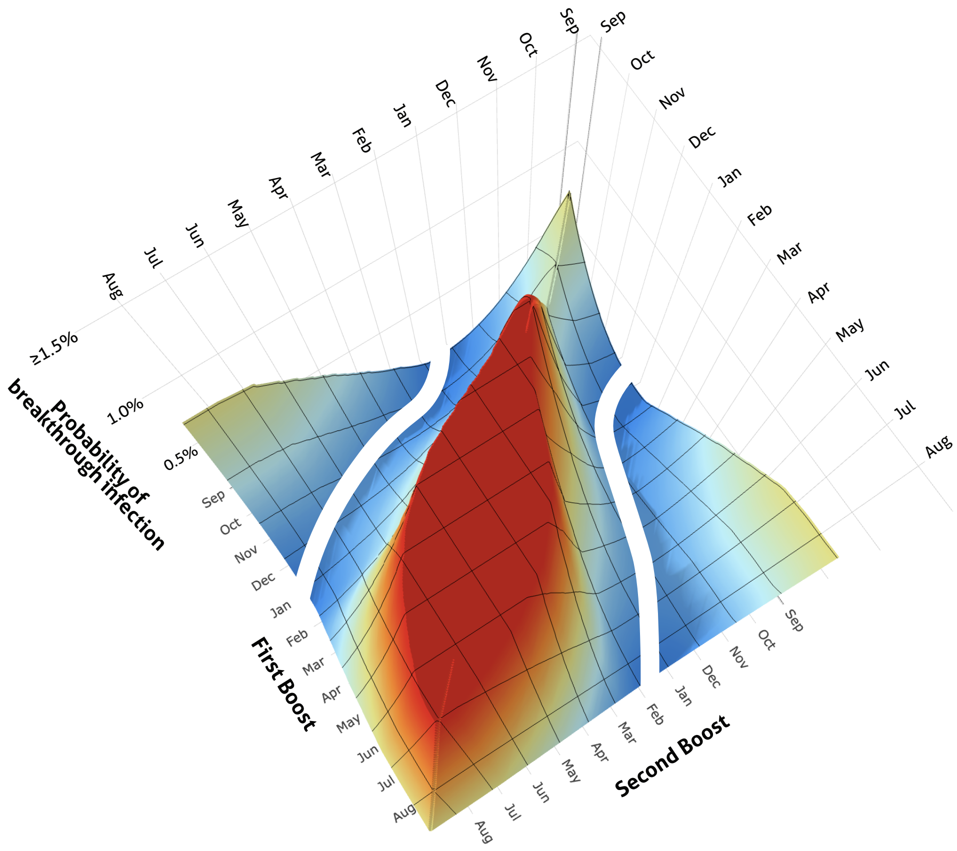


**J**

### GUANGZHOU, CHINA

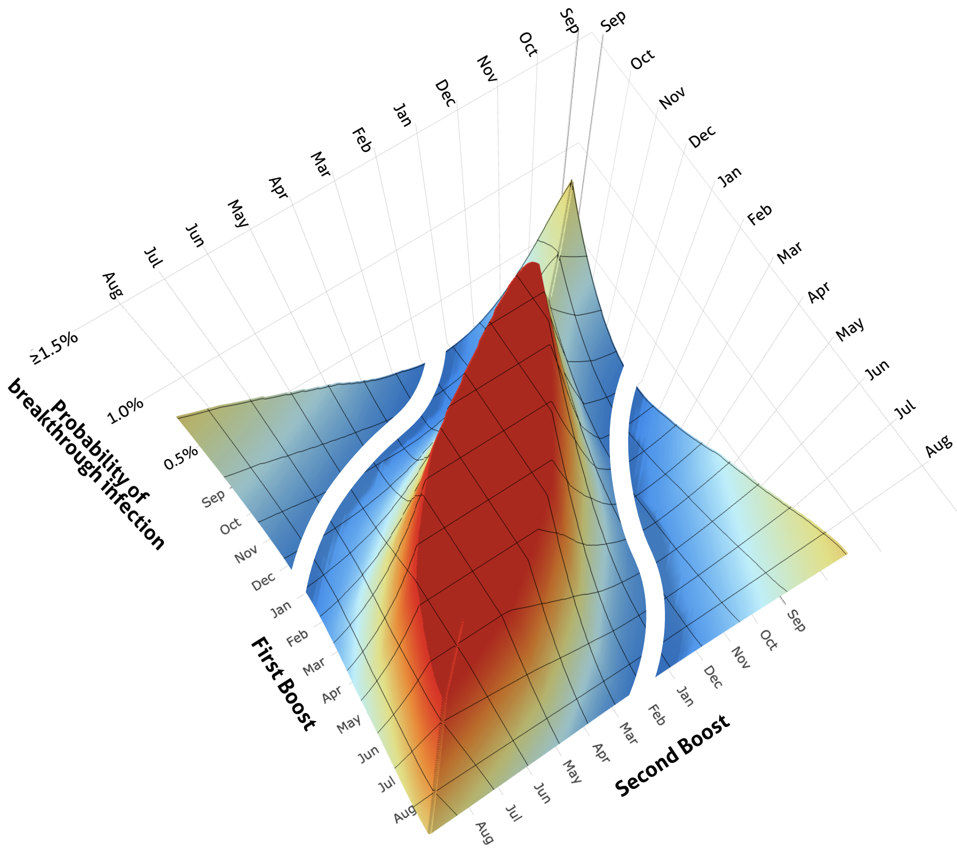

K

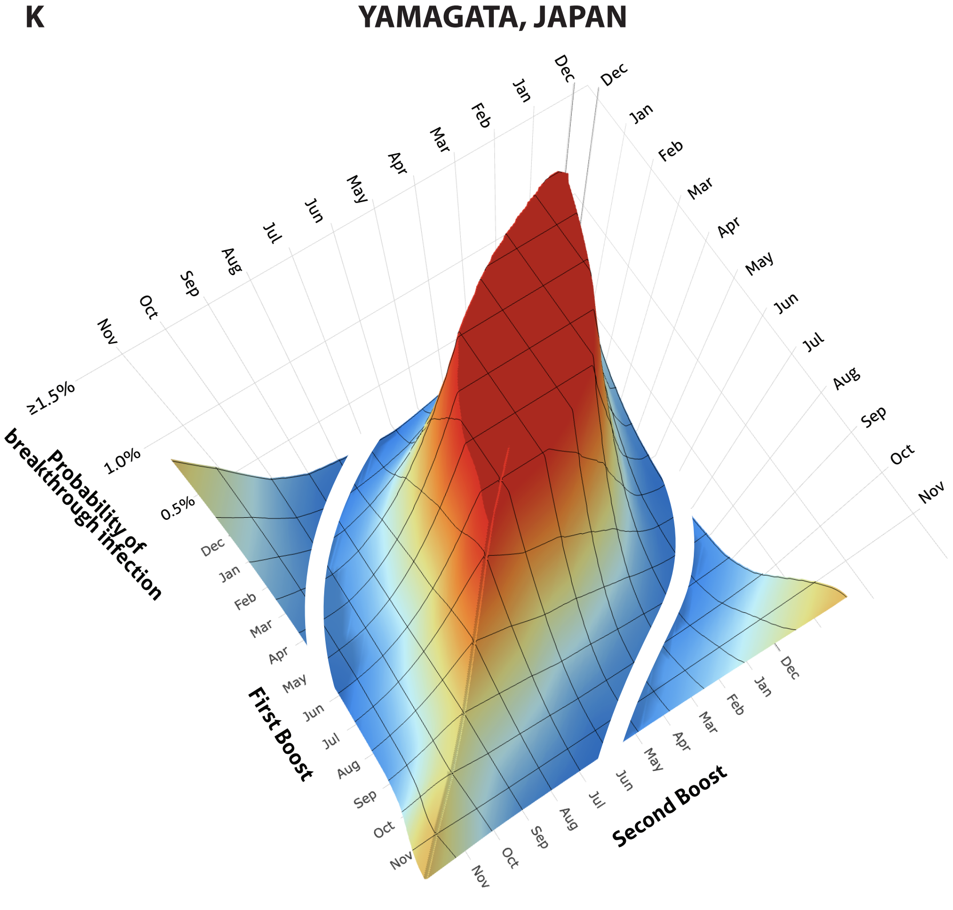


**Figure S1.** Probabilities of breakthrough infection for each biannual boost pairing (high: red, low: blue; optimal: white) for **A.** Stockholm, Sweden; **B.** Gothenburg, Sweden; **C.** Nepal; **D.** Beersheeba, Israel; **E.** New York City in 2024; **F.** Trøndelag, Norway; **G.** Amsterdam, Netherlands; **H.** South Korea; **I.** Edingburgh, UK; **J.** Guangzhou, China; **K.** Yamagata, Japan.
