## Supplementary Fig. 2 for "Optimal biannual COVID-19 vaccine boosting dates for those aged 65 and over"

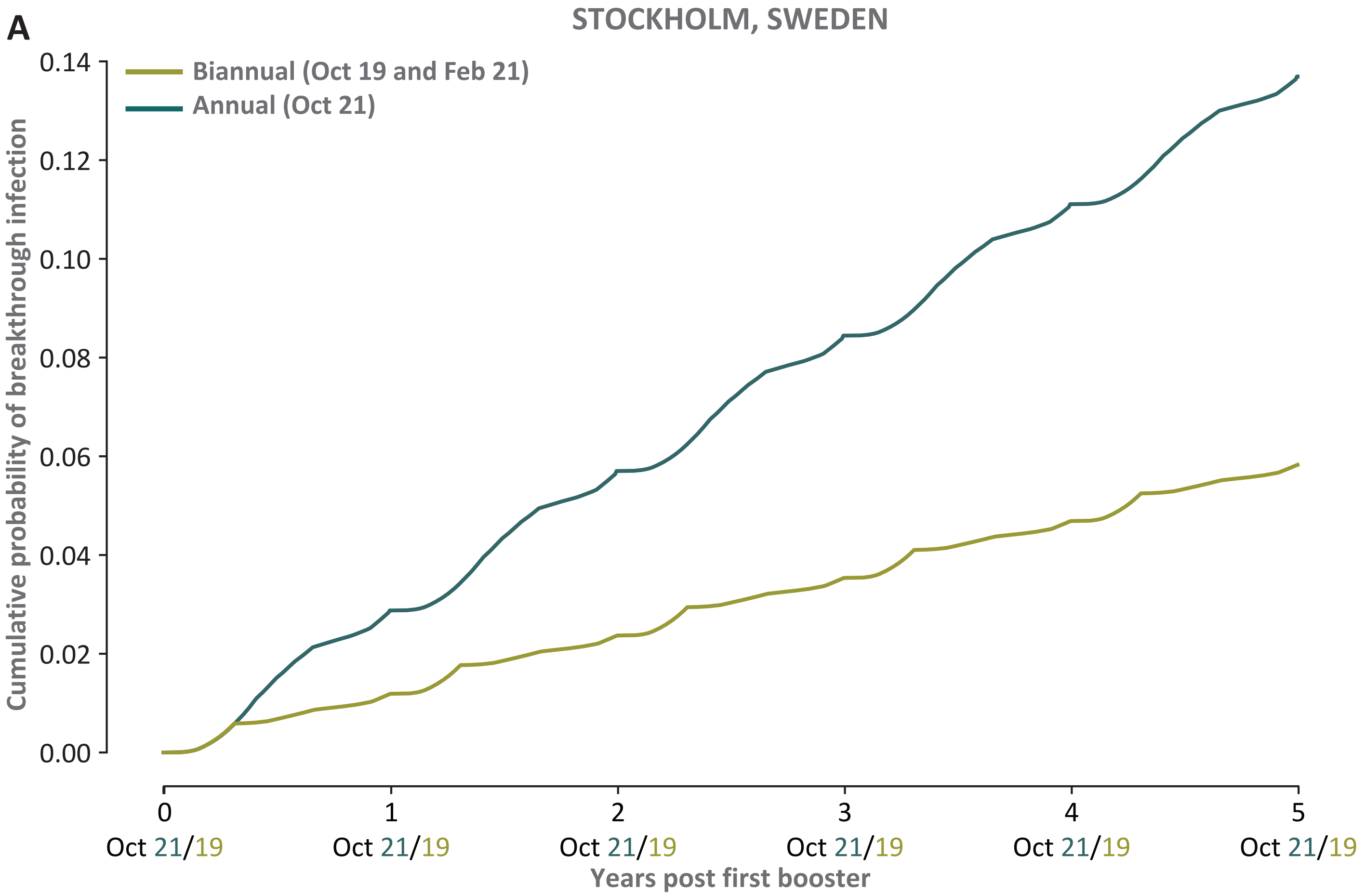
A
STOCKHOLM, SWEDEN
Biannual (Oct 19 and Feb 21)
Annual (Oct 21)
Cumulative probability of breakthrough infection
0.14
0.12
0.10
0.08
0.06
0.04
0.02
0.00
0
1
2
3
4
5
Oct 21/19
Oct 21/19
Oct 21/19
Oct 21/19
Oct 21/19
Oct 21/19
Years post first booster

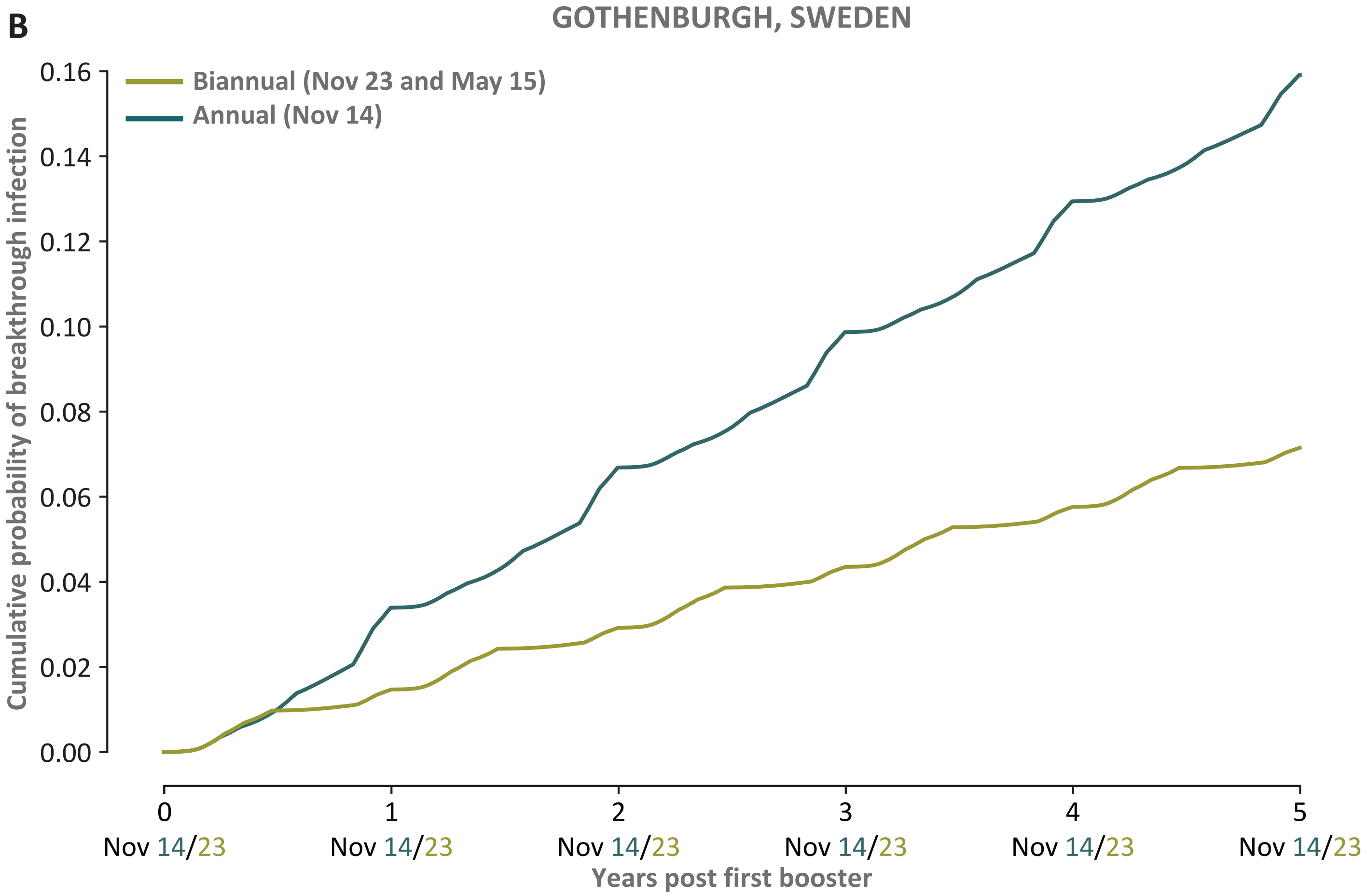
B
GOTHENBURGH, SWEDEN
Biannual (Nov 23 and May 15)
Annual (Nov 14)
Cumulative probability of breakthrough infection
0.16
0.14
0.12
0.10
0.08
0.06
0.04
0.02
0.00
0
1
2
3
4
5
Nov 14/23
Nov 14/23
Nov 14/23
Nov 14/23
Nov 14/23
Nov 14/23
Years post first booster

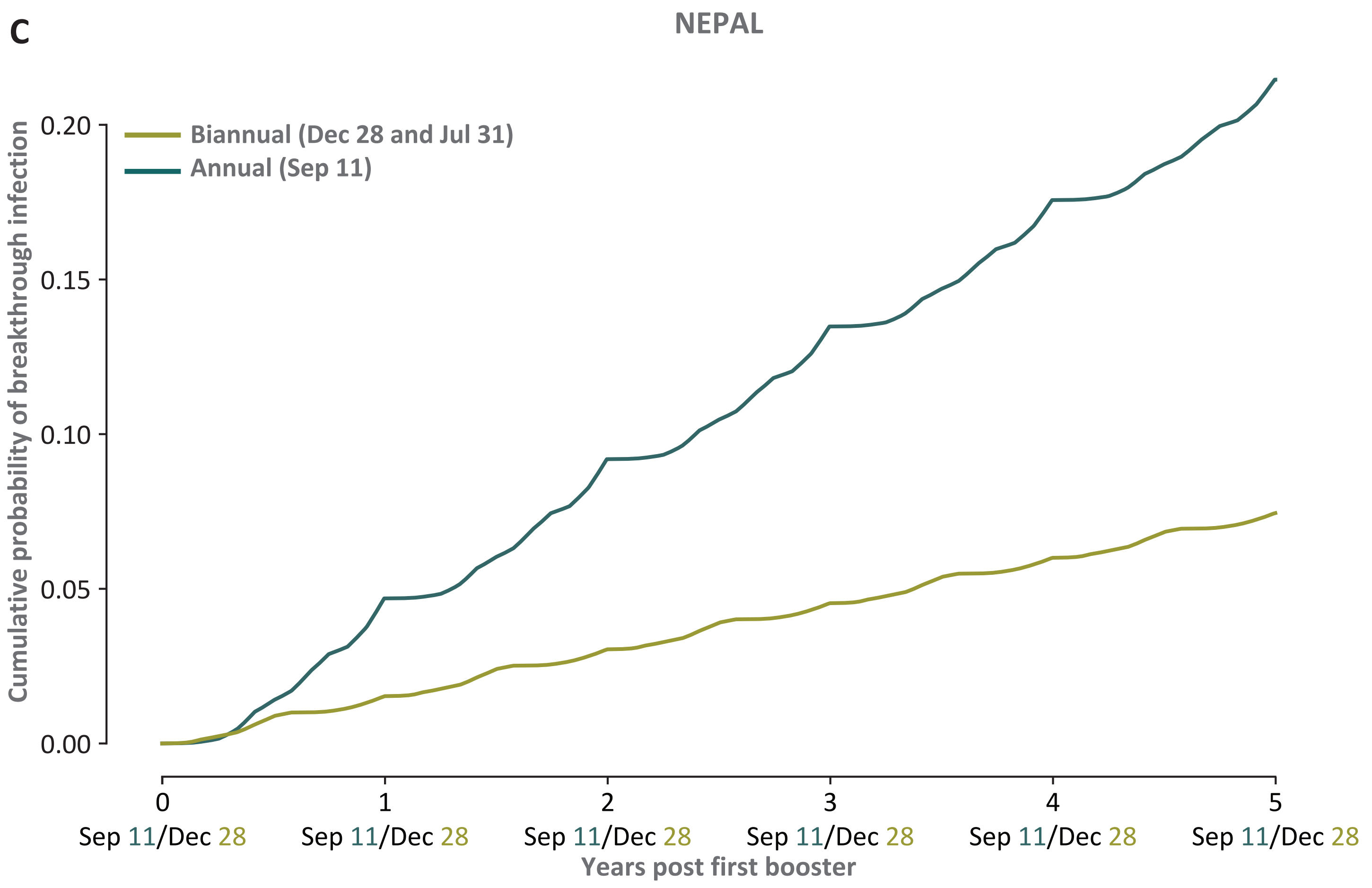
C
NEPAL
Biannual (Dec 28 and Jul 31)
Annual (Sep 11)
Cumulative probability of breakthrough infection
0.00
0.05
0.10
0.15
0.20
0
1
2
3
4
5
Sep 11/Dec 28
Sep 11/Dec 28
Sep 11/Dec 28
Sep 11/Dec 28
Sep 11/Dec 28
Sep 11/Dec 28
Years post first booster

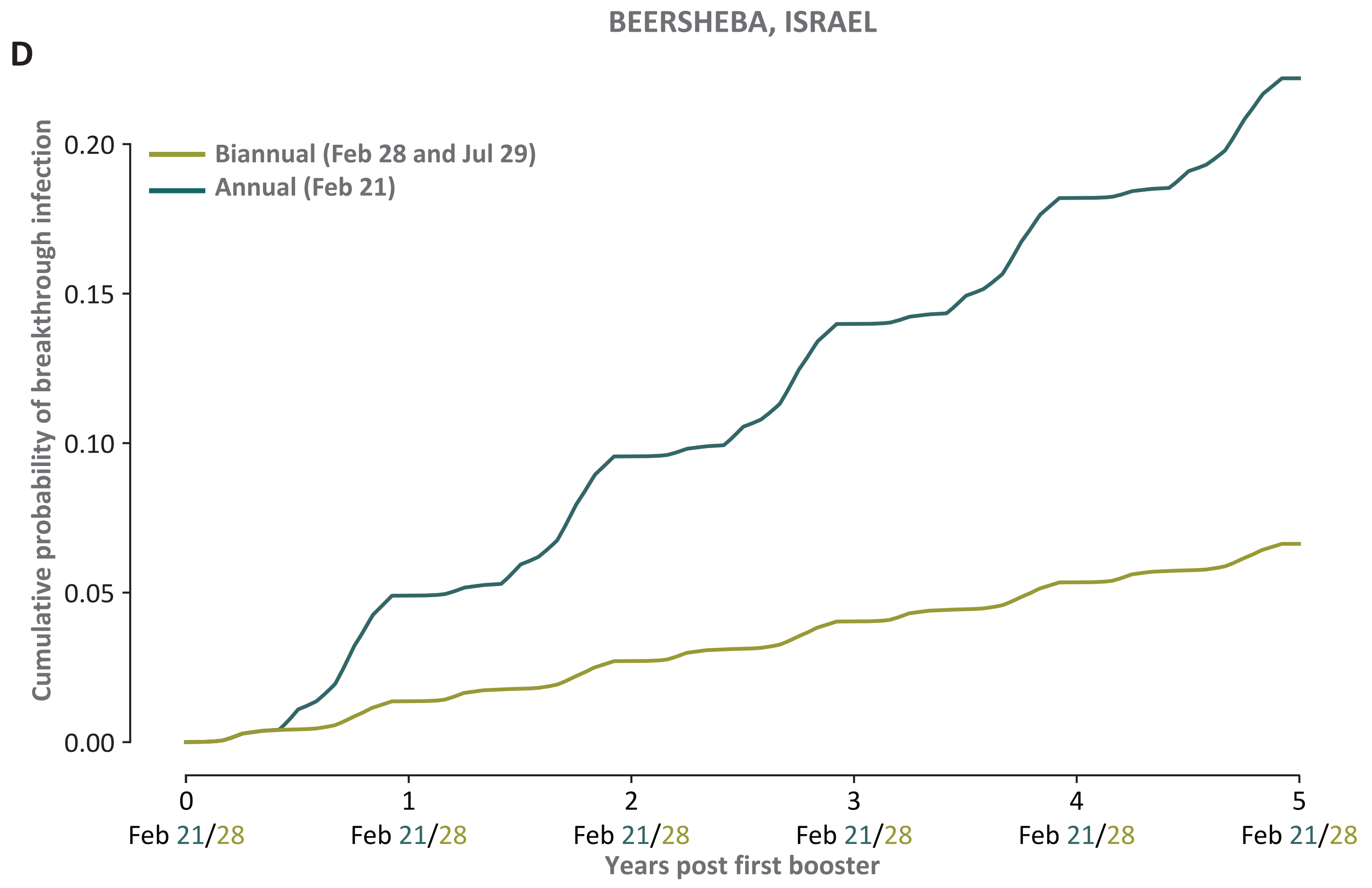
D
BEERSHEBA, ISRAEL
Biannual (Feb 28 and Jul 29)
Annual (Feb 21)
Cumulative probability of breakthrough infection
0.00
0.05
0.10
0.15
0.20
0
1
2
3
4
5
Feb 21/28
Feb 21/28
Feb 21/28
Feb 21/28
Feb 21/28
Feb 21/28
Years post first booster

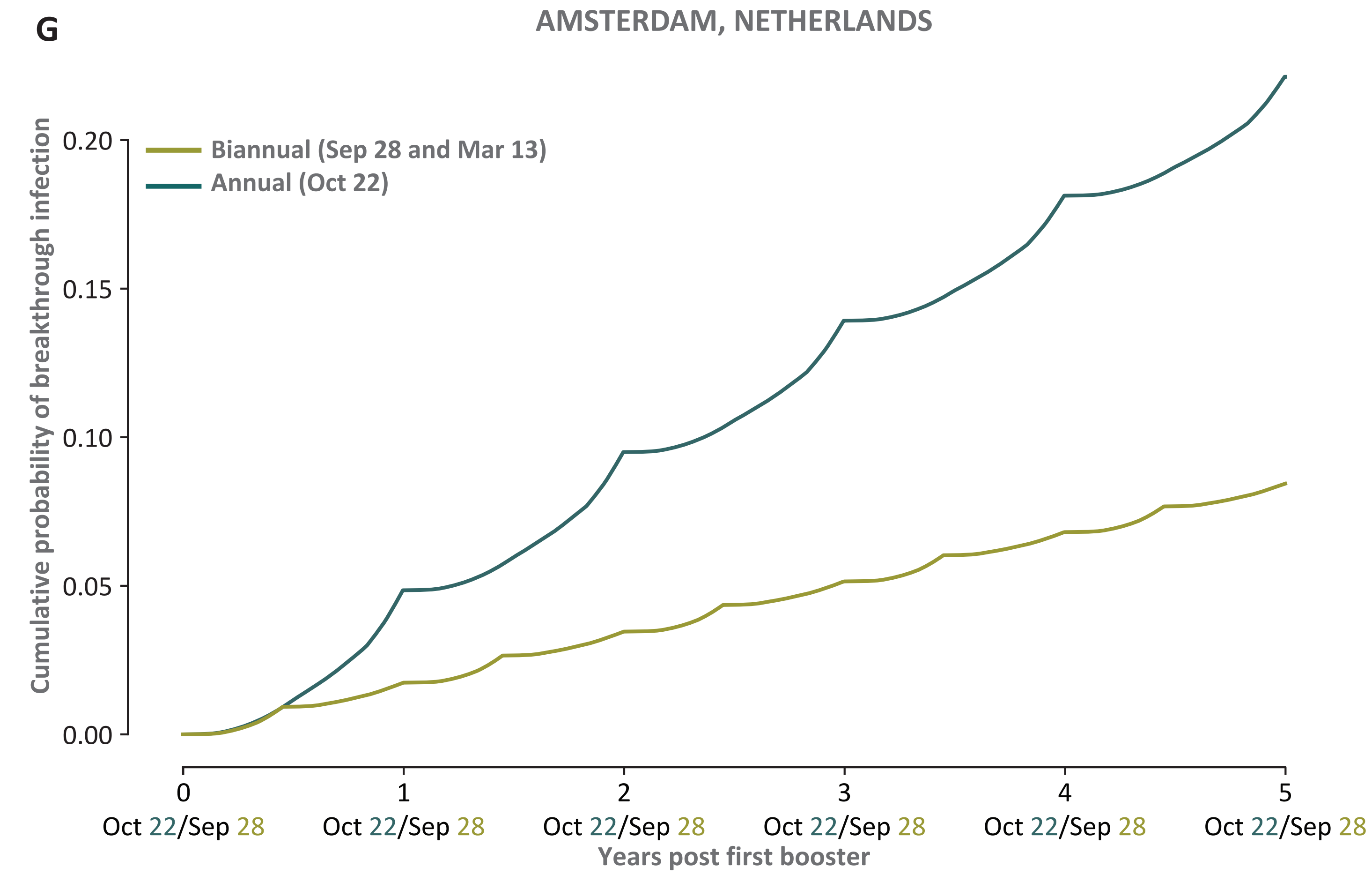
G
AMSTERDAM, NETHERLANDS
Biannual (Sep 28 and Mar 13)
Annual (Oct 22)
Cumulative probability of breakthrough infection
0.20
0.15
0.10
0.05
0.00
0
1
2
3
4
5
Oct 22/Sep 28
Oct 22/Sep 28
Oct 22/Sep 28
Oct 22/Sep 28
Oct 22/Sep 28
Oct 22/Sep 28
Years post first booster

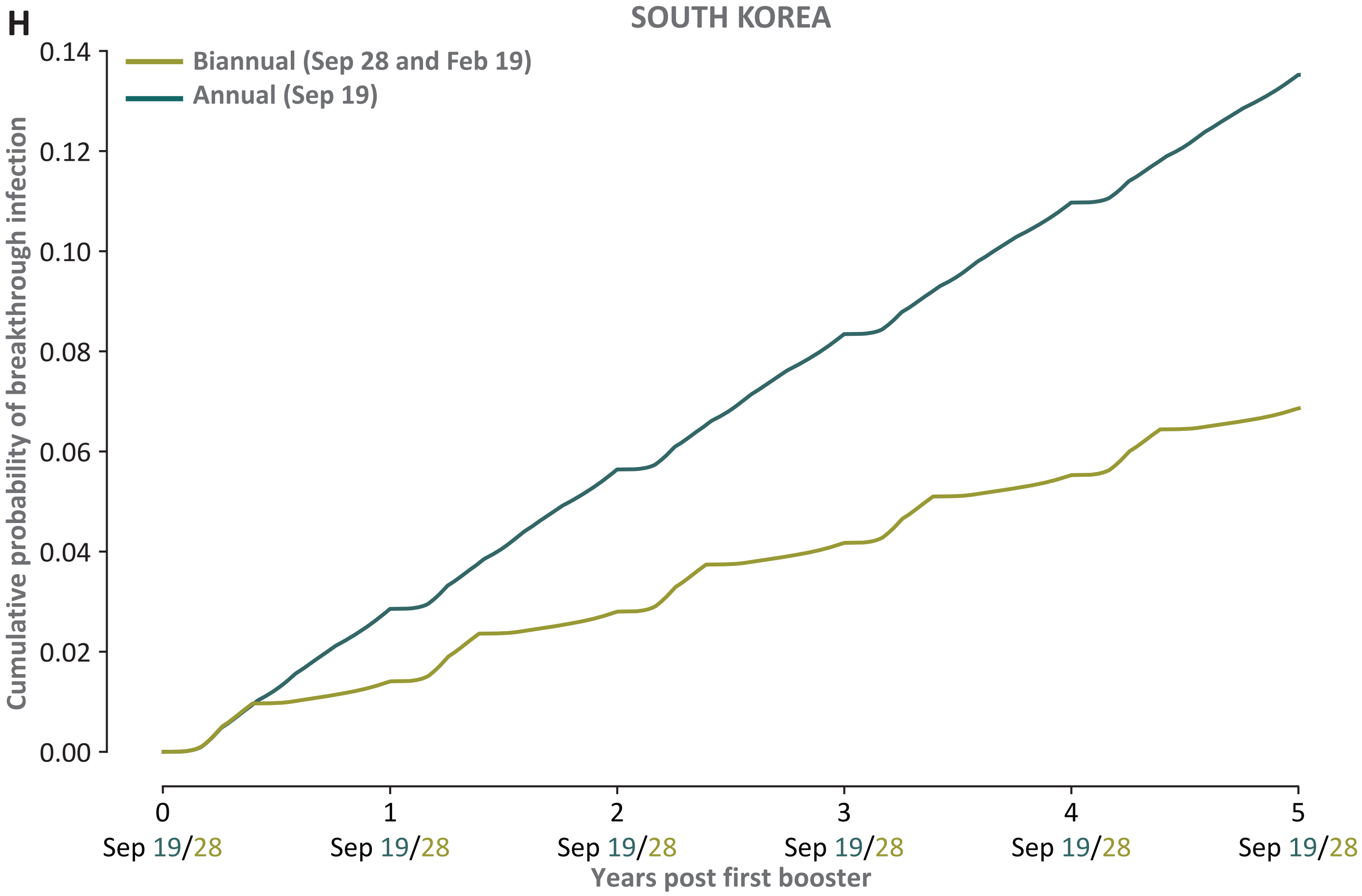
H
SOUTH KOREA
Biannual (Sep 28 and Feb 19)
Annual (Sep 19)
Cumulative probability of breakthrough infection
0.14
0.12
0.10
0.08
0.06
0.04
0.02
0.00
0
1
2
3
4
5
Sep 19/28
Sep 19/28
Sep 19/28
Sep 19/28
Sep 19/28
Sep 19/28
Years post first booster

E

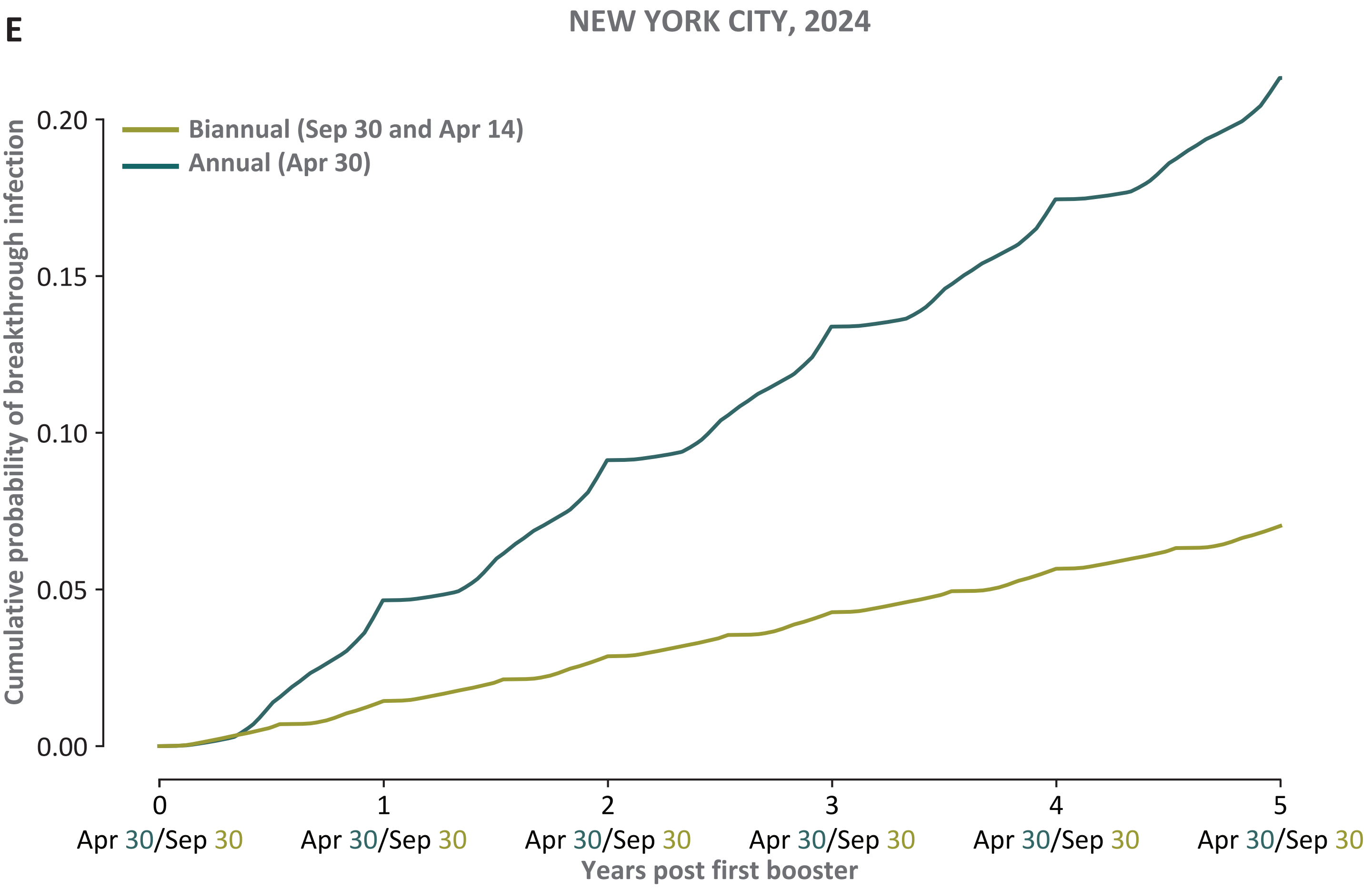
NEW YORK CITY, 2024
Biannual (Sep 30 and Apr 14)
Annual (Apr 30)
Cumulative probability of breakthrough infection
0.20
0.15
0.10
0.05
0.00
0
1
2
3
4
5
Apr 30/Sep 30
Apr 30/Sep 30
Apr 30/Sep 30
Apr 30/Sep 30
Apr 30/Sep 30
Apr 30/Sep 30
Years post first booster

F

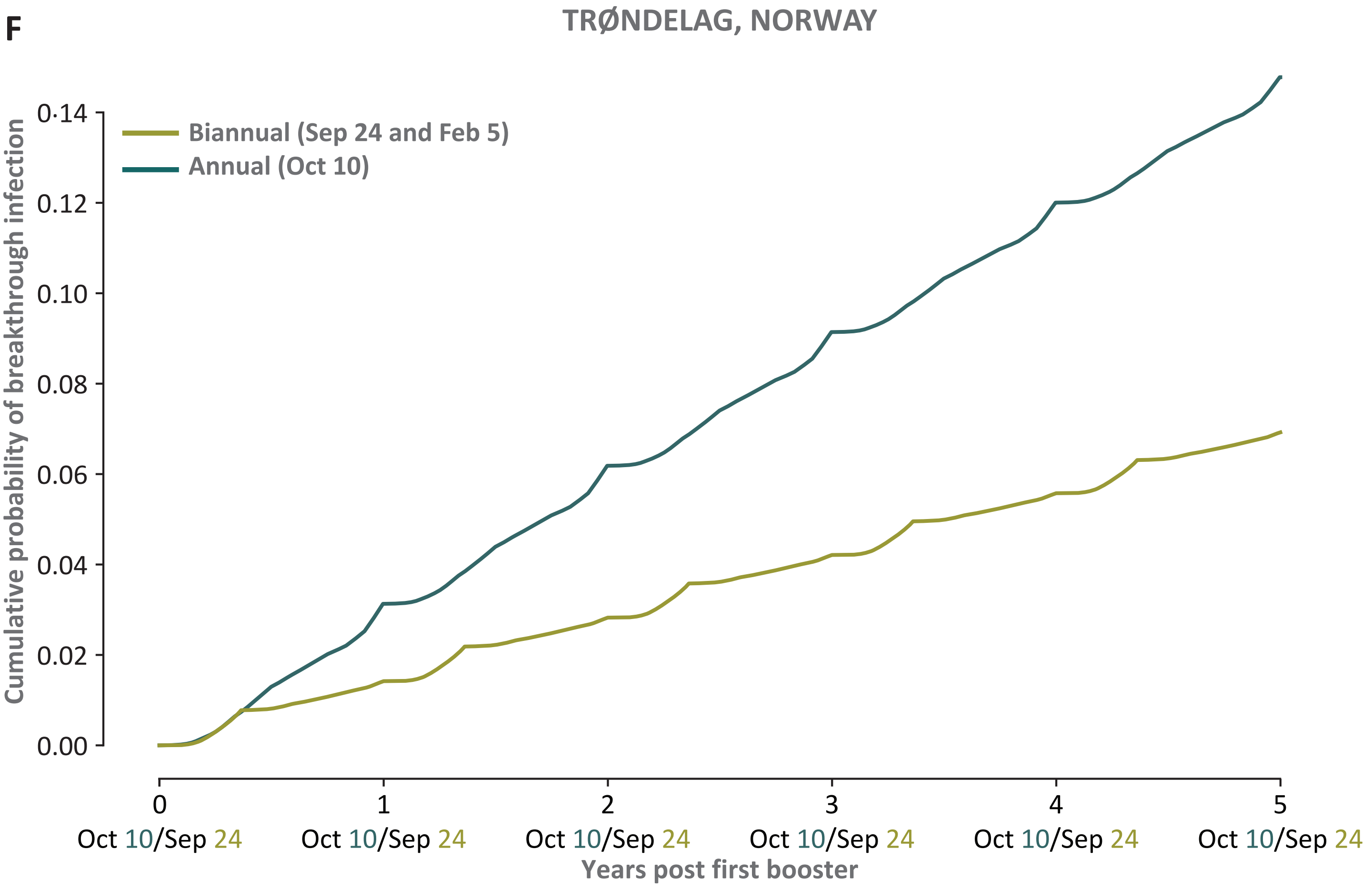
TRØNDELAG, NORWAY
Biannual (Sep 24 and Feb 5)
Annual (Oct 10)
Cumulative probability of breakthrough infection
0·14
0.12
0.10
0.08
0.06
0.04
0.02
0.00
0
1
2
3
4
5
Oct 10/Sep 24
Oct 10/Sep 24
Oct 10/Sep 24
Oct 10/Sep 24
Oct 10/Sep 24
Oct 10/Sep 24
Years post first booster

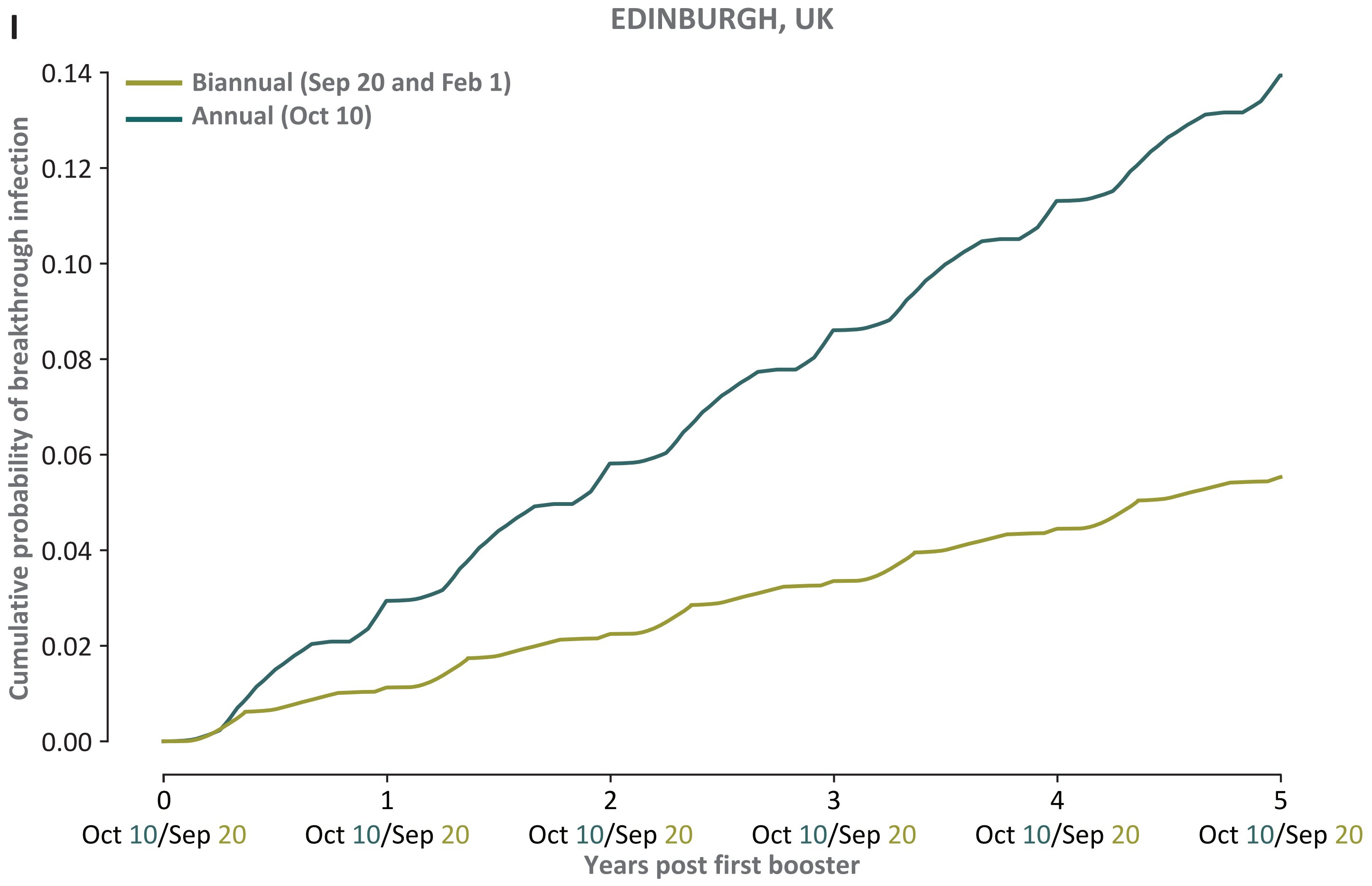
I
EDINBURGH, UK
Biannual (Sep 20 and Feb 1)
Annual (Oct 10)
Cumulative probability of breakthrough infection
0.14
0.12
0.10
0.08
0.06
0.04
0.02
0.00
0
1
2
3
4
5
Oct 10/Sep 20
Oct 10/Sep 20
Oct 10/Sep 20
Oct 10/Sep 20
Oct 10/Sep 20
Oct 10/Sep 20
Years post first booster

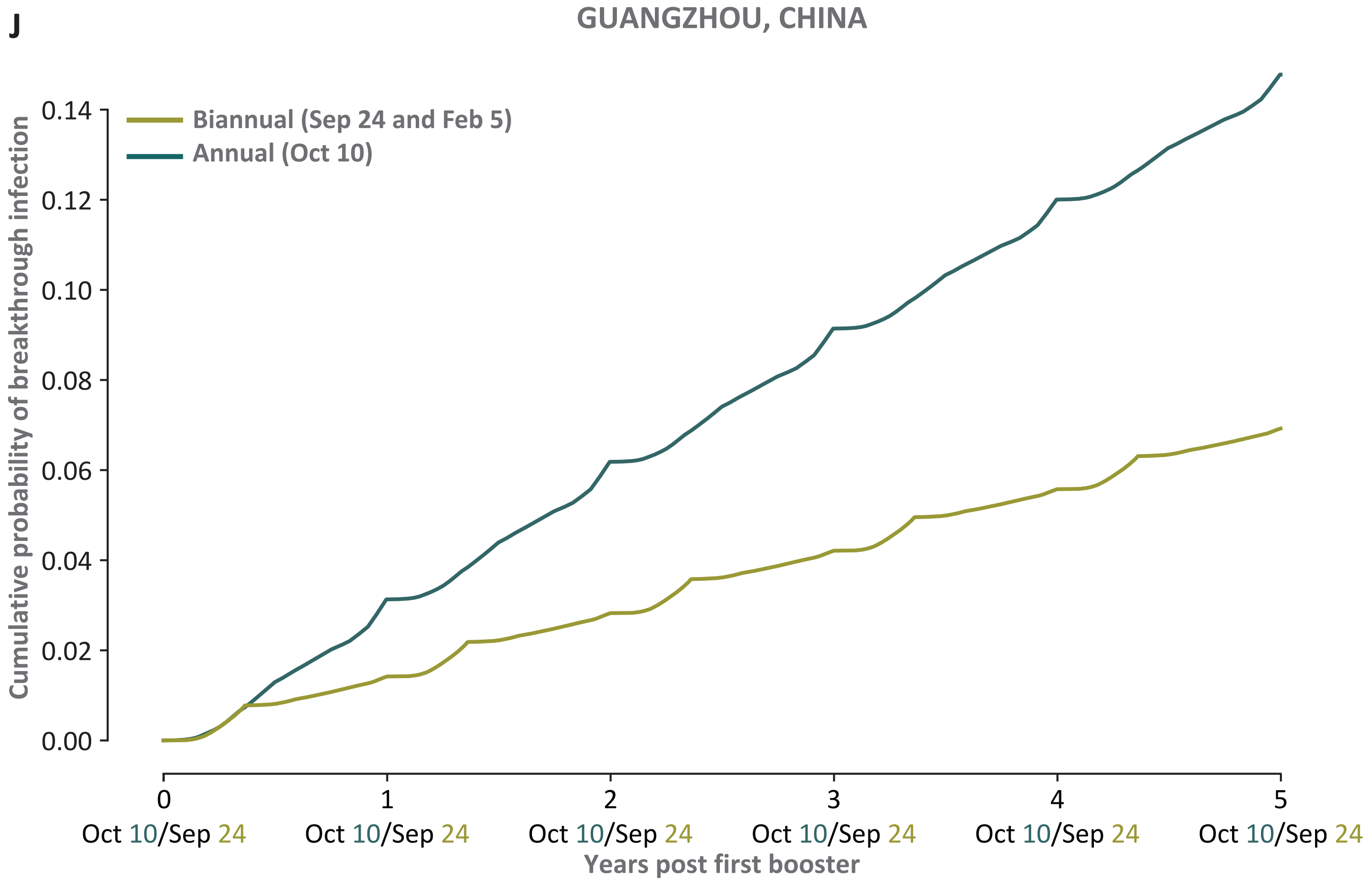
J
GUANGZHOU, CHINA
Biannual (Sep 24 and Feb 5)
Annual (Oct 10)
Cumulative probability of breakthrough infection
0.14
0.12
0.10
0.08
0.06
0.04
0.02
0.00
0
1
2
3
4
5
Oct 10/Sep 24
Oct 10/Sep 24
Oct 10/Sep 24
Oct 10/Sep 24
Oct 10/Sep 24
Oct 10/Sep 24
Years post first booster

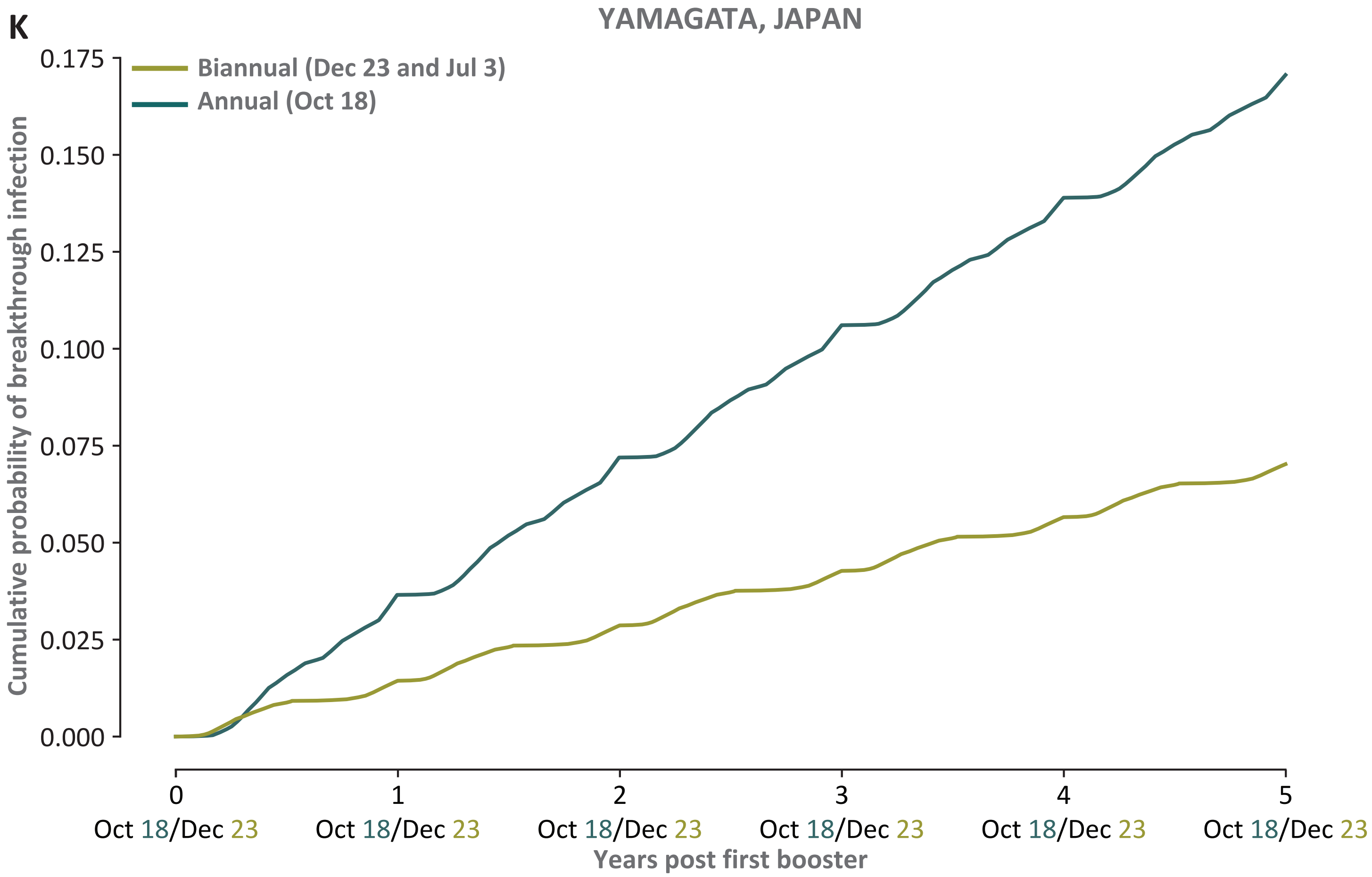


**Figure S2**. Cumulative probabilities of breakthrough infection over five years, with optimal annual boosting (dark teal) and with optimal biannual boosting (green) for **A.** Stockholm, Sweden; **B.** Gothenburg, Sweden; **C.** Nepal; **D.** Beersheeba, Israel; **E.** New York City in 2024; **F.** Trøndelag, Norway; **G.** Amsterdam, Netherlands; **H.** South Korea; **I.** Edingburgh, UK; **J.** Guangzhou, China; **K.** Yamagata, Japan.
