## Supplementary Fig. 3 for "Optimal biannual COVID-19 vaccine boosting dates for those aged 65 and over"

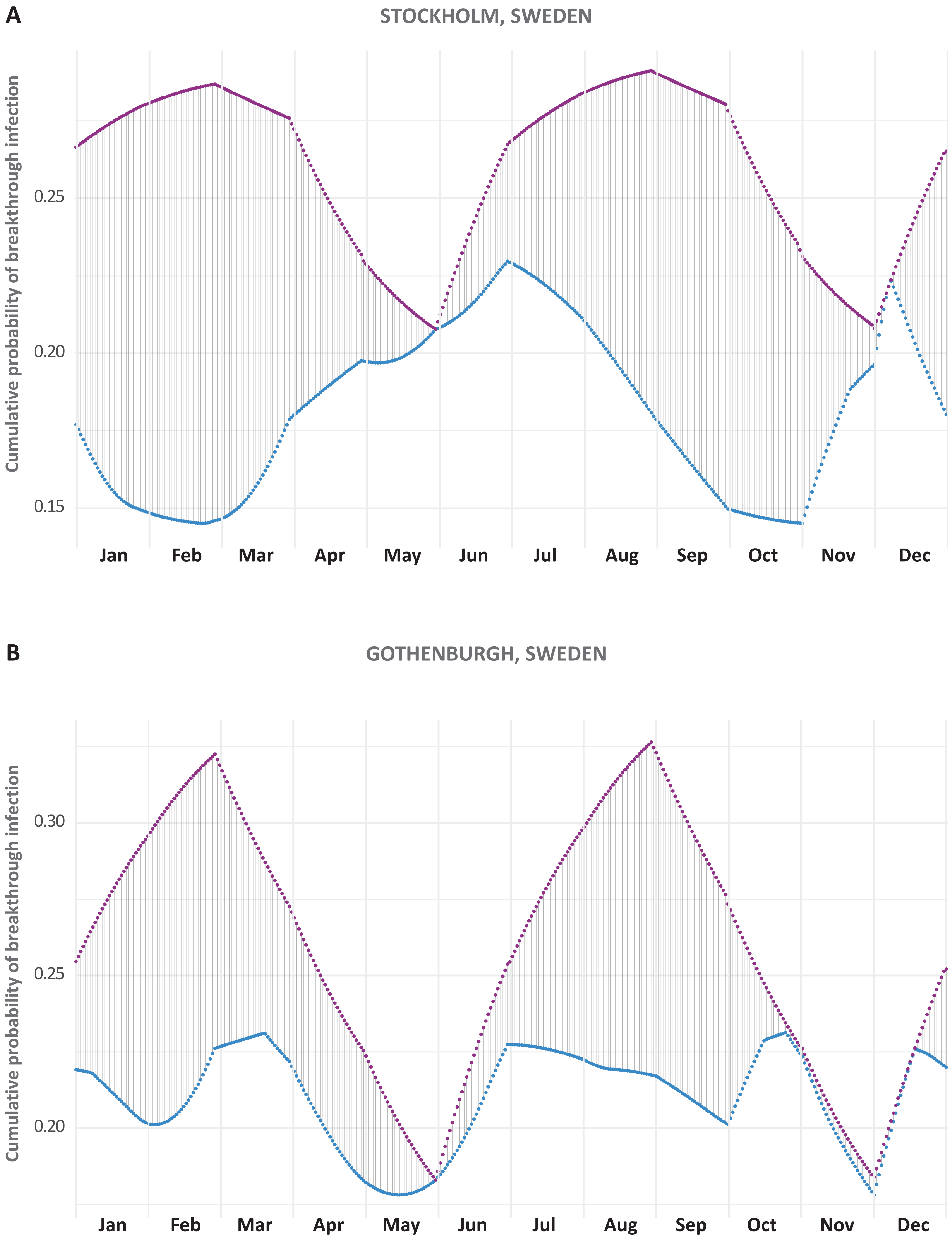
A
STOCKHOLM, SWEDEN
Cumulative probability of breakthrough infection
0.25
0.20
0.15
Jan
Feb
Mar
Apr
May
Jun
Jul
Aug
Sep
Oct
Nov
Dec
B
GOTHENBURGH, SWEDEN
Cumulative probability of breakthrough infection
0.30
0.25
0.20
Jan
Feb
Mar
Apr
May
Jun
Jul
Aug
Sep
Oct
Nov
Dec

C

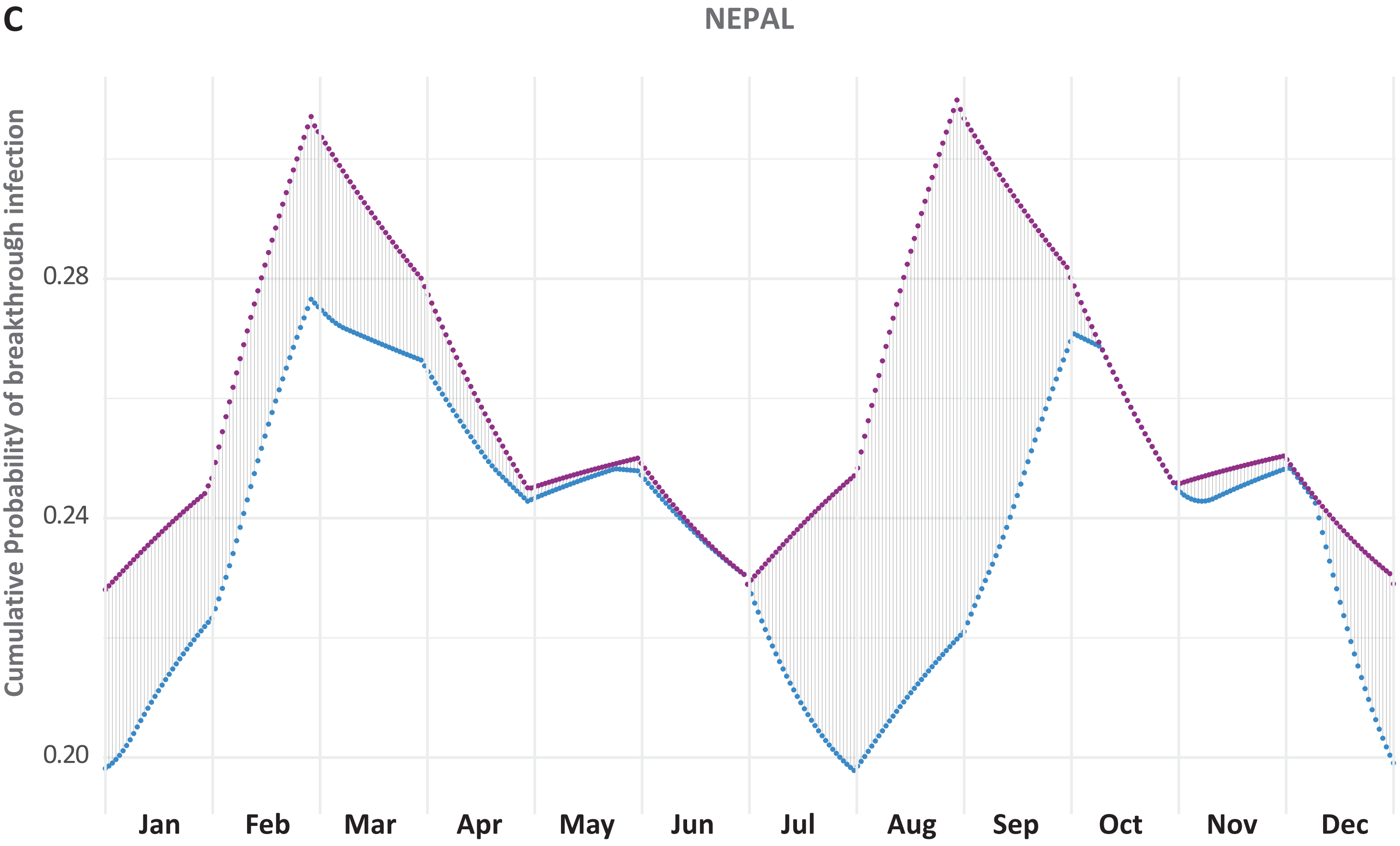
NEPAL
Cumulative ProbabilitY of breakthrough infection
0.28
0.24
0.20
Jan
Feb
Mar
Apr
May
Jun
Jul
Aug
Sep
Oct
Nov
Dec

D

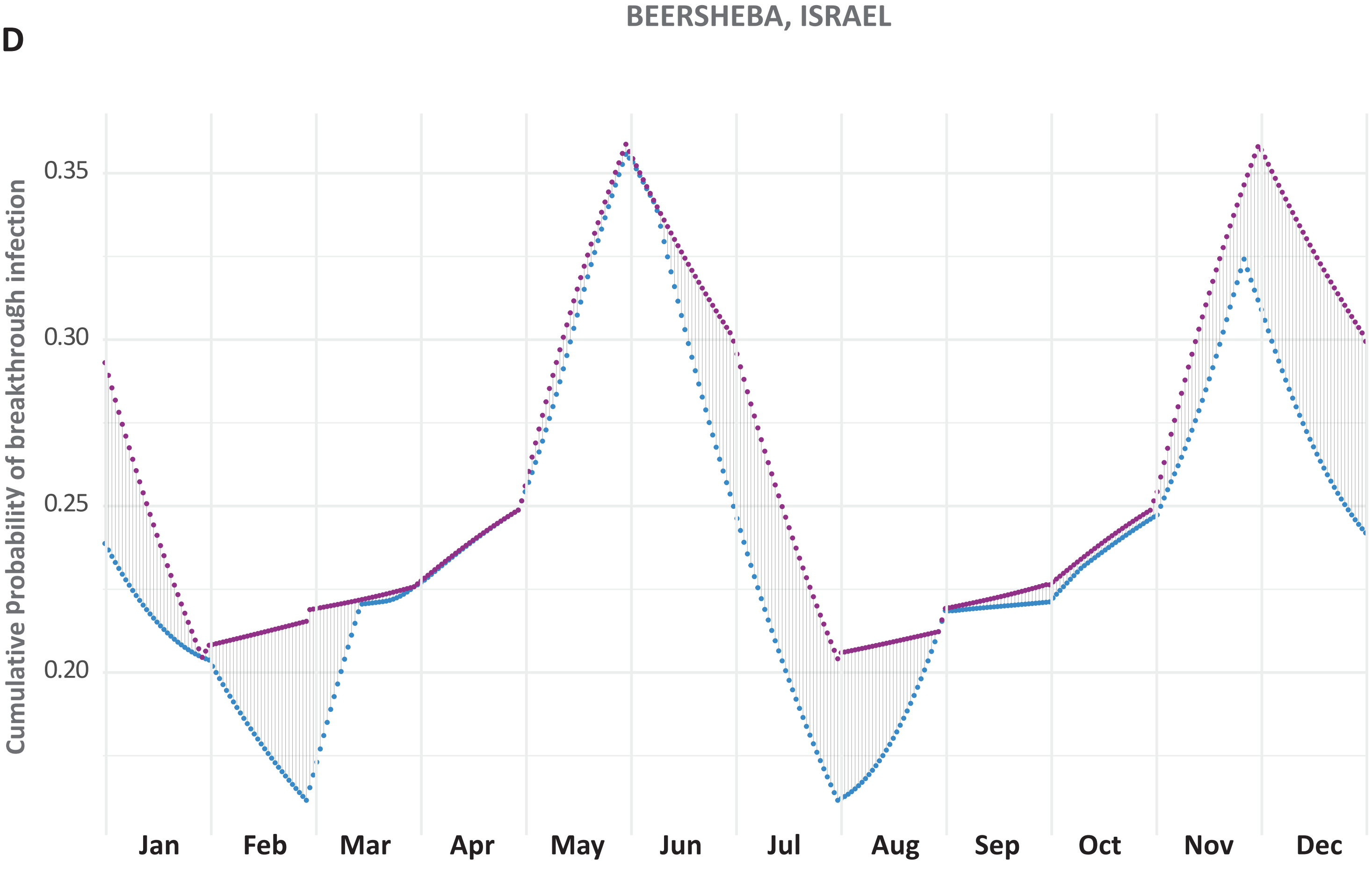
BEERSHEBA, ISRAEL
Cumulative ProbabilitY of breakthrough infection
0.35
0.30
0.25
0.20
Jan
Feb
Mar
Apr
May
Jun
Jul
Aug
Sep
Oct
Nov
Dec

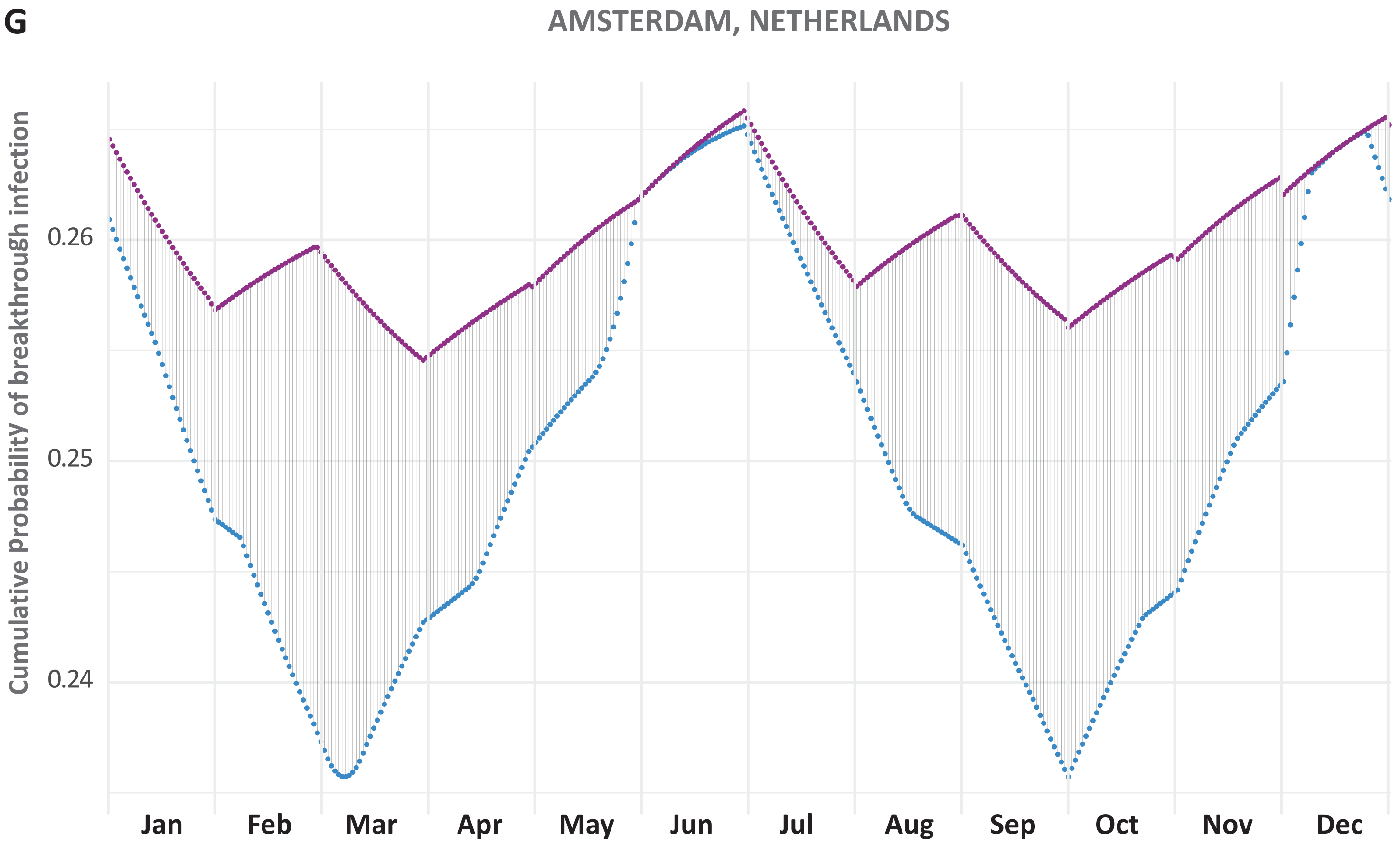
G
AMSTERDAM, NETHERLANDS
Cumulative ProbabilitY of breakthrough infection
0.26
0.25
0.24
Jan
Feb
Mar
Apr
May
Jun
Jul
Aug
Sep
Oct
Nov
Dec

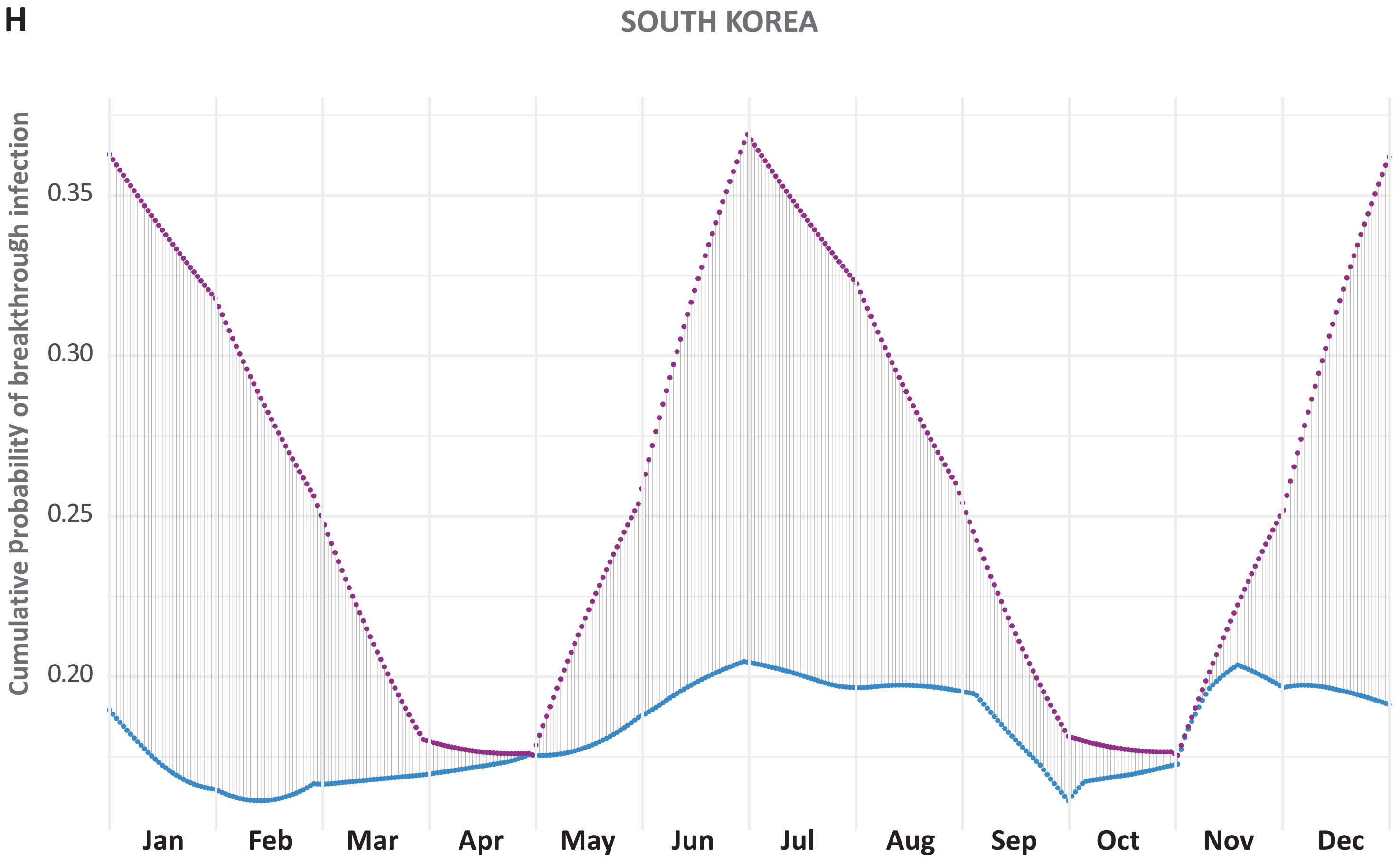
H
SOUTH KOREA
Cumulative ProbabilitY of breakthrough infection
0.35
0.30
0.25
0.20
Jan
Feb
Mar
Apr
May
Jun
Jul
Aug
Sep
Oct
Nov
Dec

**E**

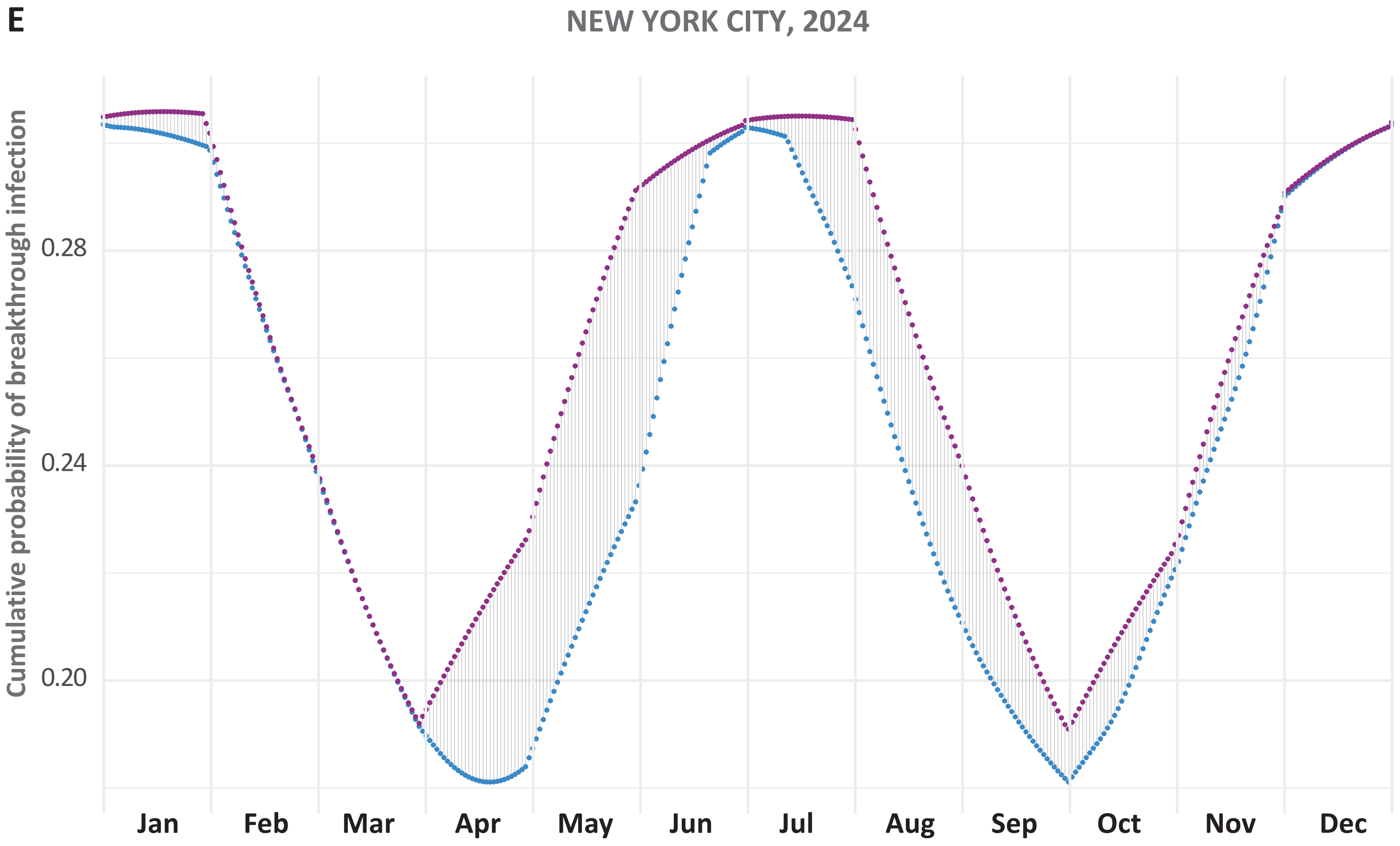
NEW YORK CITY, 2024
Cumulative probability of breakthrough infection
0.28
0.24
0.20
Jan
Feb
Mar
Apr
May
Jun
Jul
Aug
Sep
Oct
Nov
Dec

**F**

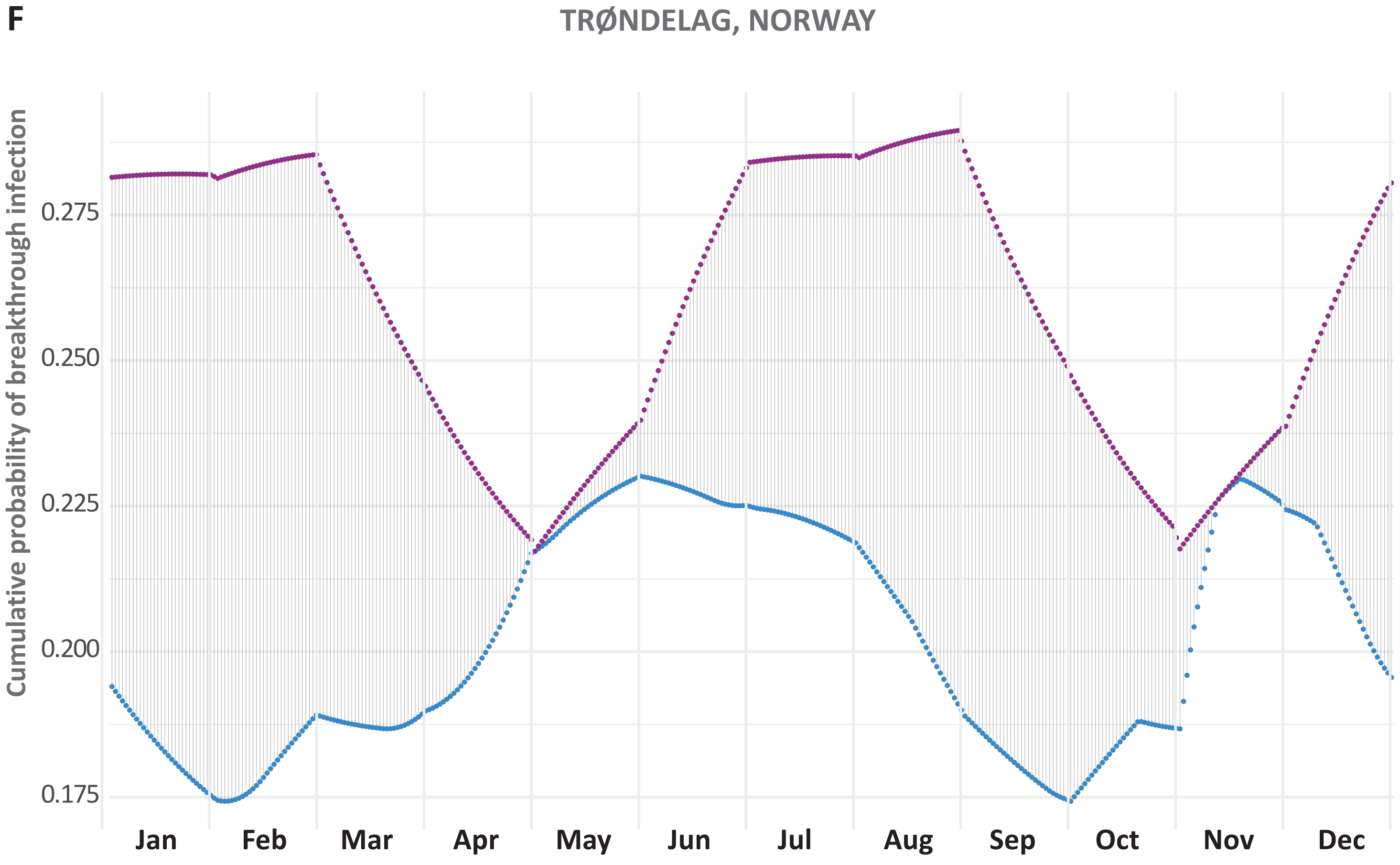
TRØNDELAG, NORWAY
Cumulative probability of breakthrough infection
0.275
0.250
0.225
0.200
0.175
Jan
Feb
Mar
Apr
May
Jun
Jul
Aug
Sep
Oct
Nov
Dec

I

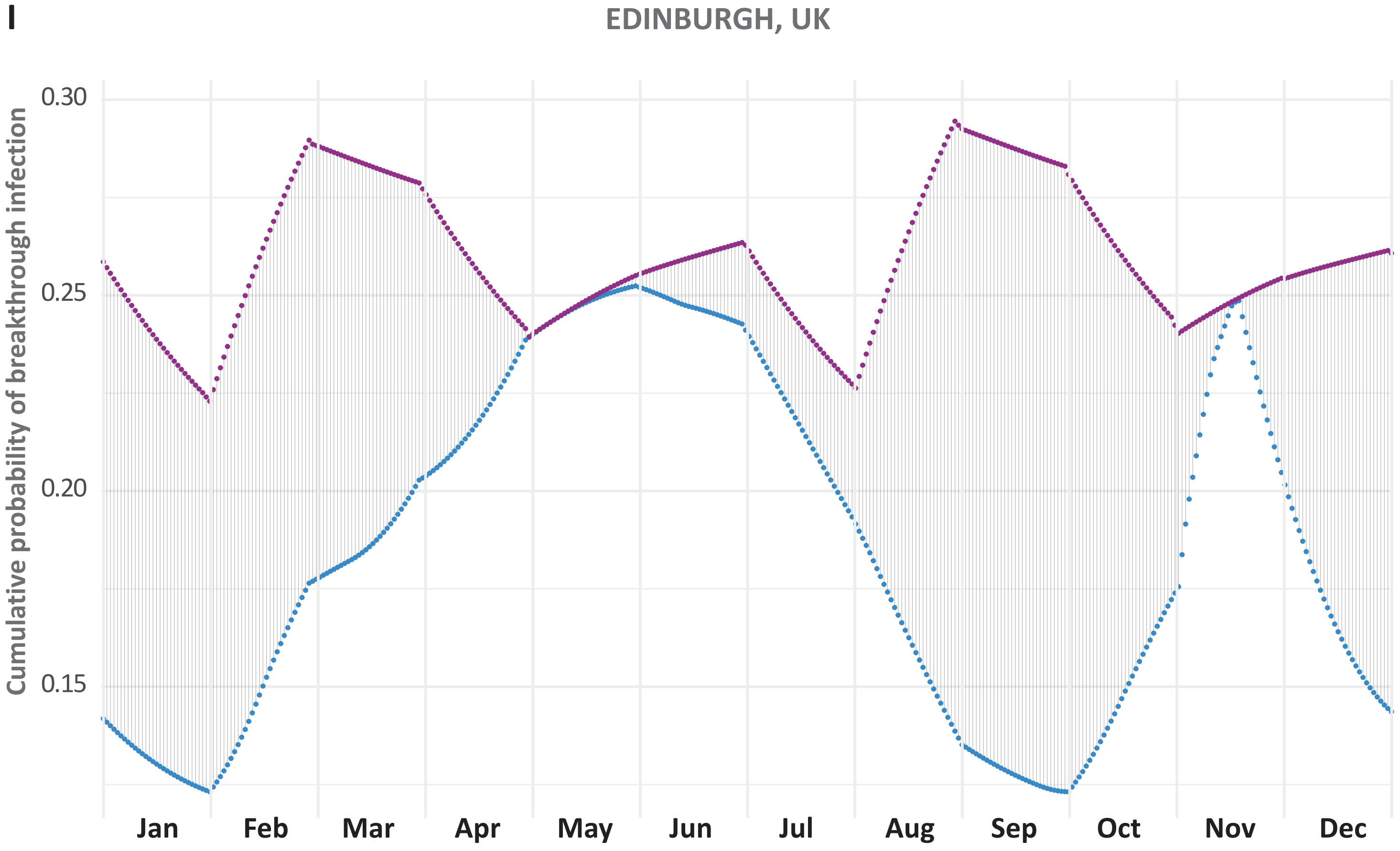

EDINBURGH, UK
Cumulative probability of breakthrough infection
0.30
0.25
0.20
0.15
Jan
Feb
Mar
Apr
May
Jun
Jul
Aug
Sep
Oct
Nov
Dec


J

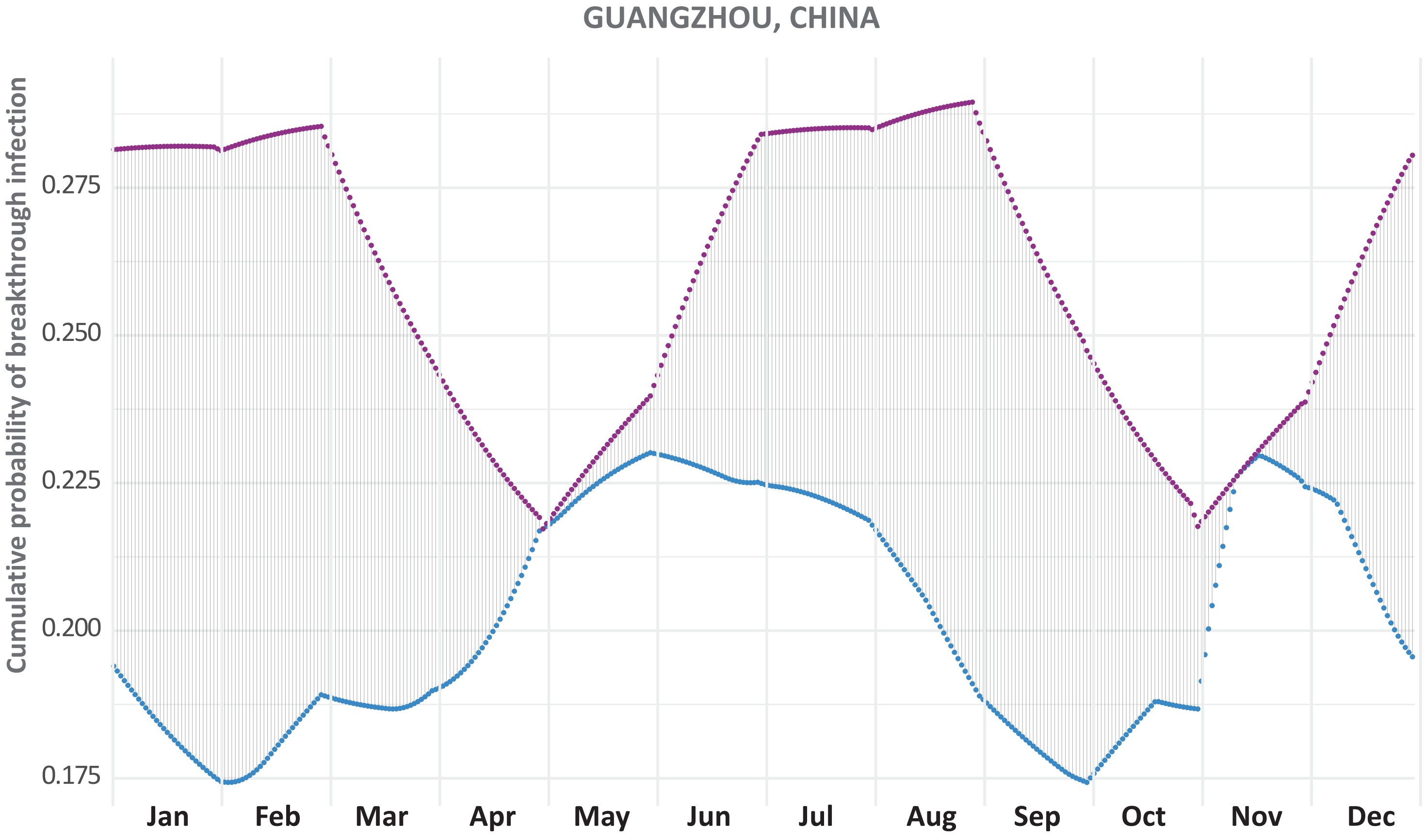

GUANGZHOU, CHINA
Cumulative probability of breakthrough infection
0.275
0.250
0.225
0.200
0.175
Jan
Feb
Mar
Apr
May
Jun
Jul
Aug
Sep
Oct
Nov
Dec

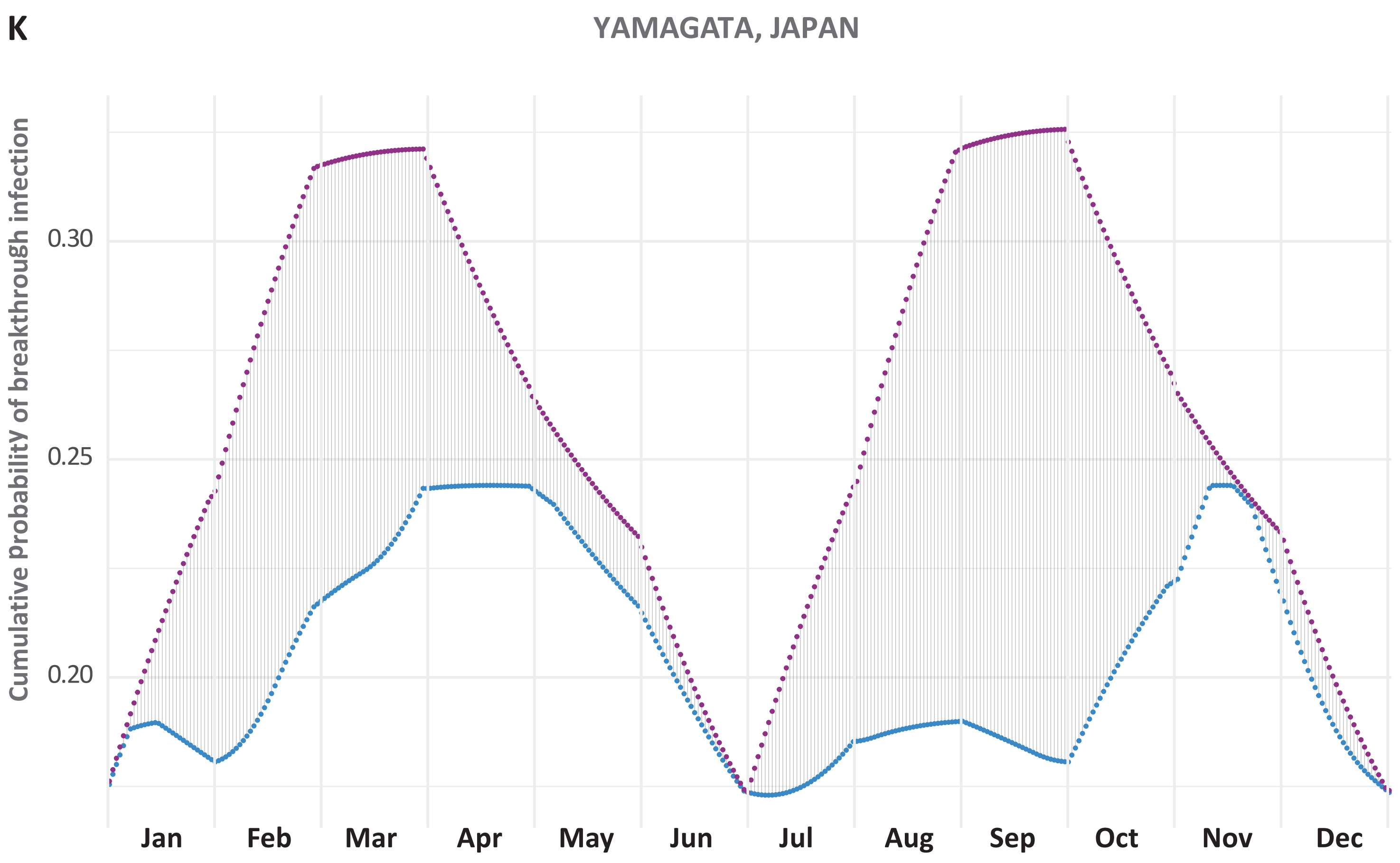
K
YAMAGATA, JAPAN
Cumulative ProbabilitY of breakthrough infection
0.30
0.25
0.20
Jan
Feb
Mar
Apr
May
Jun
Jul
Aug
Sep
Oct
Nov
Dec

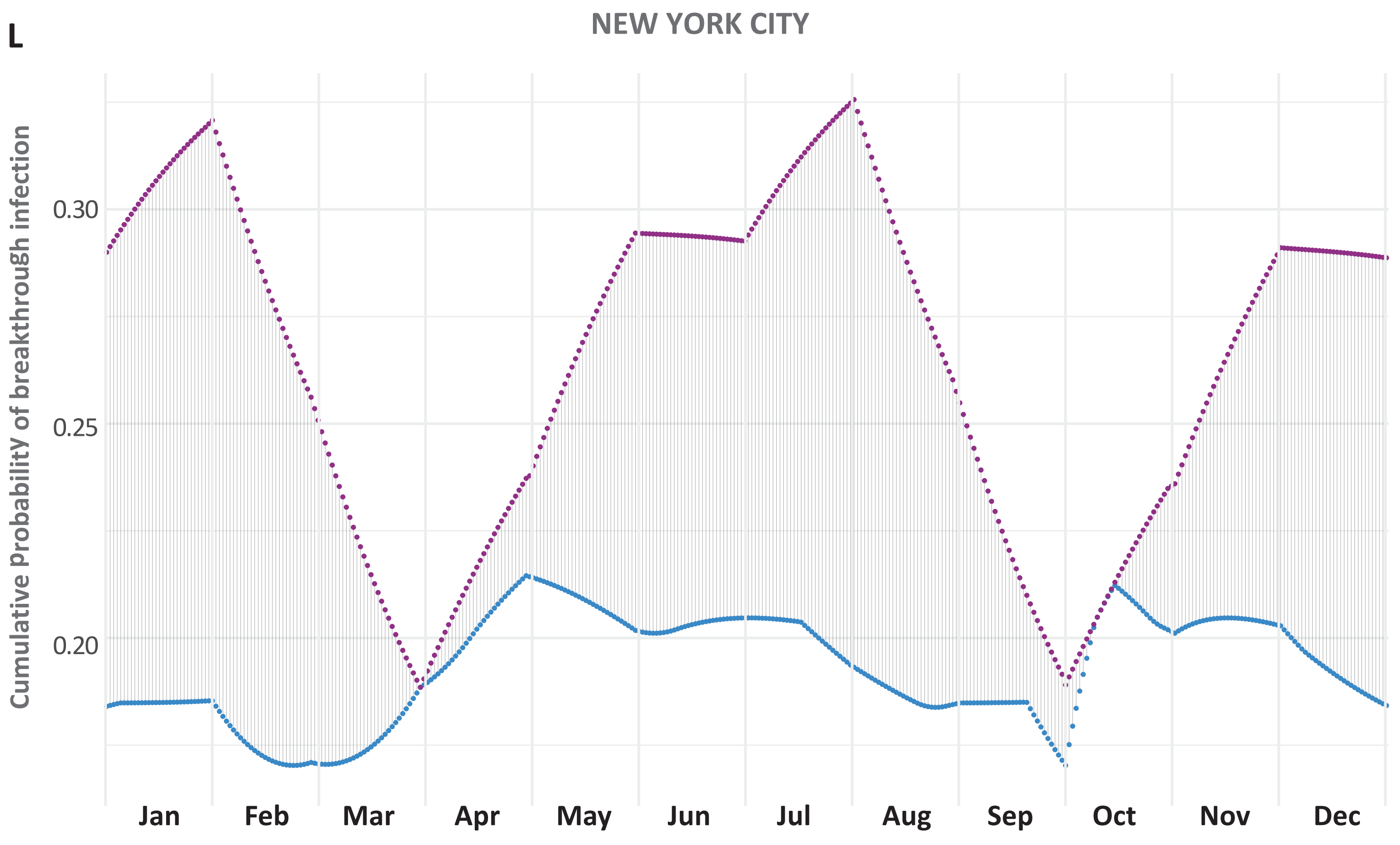
L
NEW YORK CITY
Cumulative ProbabilitY of breakthrough infection
0.30
0.25
0.20
Jan
Feb
Mar
Apr
May
Jun
Jul
Aug
Sep
Oct
Nov
Dec

**Figure S3**. Cumulative probabilities of breakthrough infection over one year for optimal biannual boost pairings (blue) and six month booster pairings (purple) for **A.** Stockholm, Sweden; **B.** Gothenburg, Sweden; **C.** Nepal; **D.** Beersheeba, Israel; **E.** New York City in 2024; **F.** Trøndelag, Norway; **G.** Amsterdam, Netherlands; **H.** South Korea; **I.** Edingburgh, UK; **J.** Guangzhou, China; **K.** Yamagata, Japan, **L.** New York City, New York.
